# Trio-based whole exome sequencing in patients with suspected sporadic inborn errors of immunity: a retrospective cohort study

**DOI:** 10.1101/2022.03.30.22272929

**Authors:** Anne Hebert, Annet Simons, Janneke H.M. Schuurs-Hoeijmakers, Hans J.P.M. Koenen, Evelien Zonneveld-Huijssoon, Stefanie S.V. Henriet, Ellen J.H. Schatorjé, Esther P.A.H. Hoppenreijs, Erika K.S.M. Leenders, Etienne J.M. Janssen, Gijs W.E. Santen, Sonja A. de Munnik, Simon V. van Reijmersdal, Esther van Rijssen, Simone Kersten, Mihai G. Netea, Ruben L. Smeets, Frank L. van de Veerdonk, Alexander Hoischen, Caspar I. van der Made

**Author notes:** **Corresponding author:** Name: Alexander Hoischen, Ph.D., Address: Department of Internal Medicine and Department of Human Genetics, Radboud Center for Infectious Diseases (RCI), Radboud Institute of Molecular Life Sciences (RIMLS), Radboud University Medical Center, Nijmegen 6525GA, The Netherlands.nr.

## Abstract

**Background:** D*e novo* variants (DNVs) are currently not routinely evaluated as part of diagnostic whole exome sequencing (WES) analysis in patients with suspected inborn errors of immunity (IEI).

**Methods:** This study explored the potential added value of systematic assessment of DNVs in a retrospective cohort of 123 patients with a suspected sporadic IEI who underwent patient-parent trio-based WES.

**Results:** A likely molecular diagnosis for (part) of the immunological phenotype was achieved in 12 patients with the diagnostic *in silico* IEI WES gene panel. Exome-wide evaluation of rare, non-synonymous DNVs affecting coding or splice site regions led to the identification of 14 candidate DNVs in genes with an annotated immune function. DNVs were identified in IEI genes (*NLRP3* and *RELA*) and potentially novel candidate genes, including *PSMB10*, *DDX1*, *KMT2C* and *FBXW11*. The *FBXW11* canonical splice site DNV, in a patient with autoinflammatory disease, was shown to lead to defective RNA splicing, increased NF-κB p65 signalling, and elevated IL-1β production in primary immune cells.

**Conclusions:** This retrospective cohort study advocates the implementation of trio-based sequencing in routine diagnostics of patients with sporadic IEI. Furthermore, we have provided functional evidence supporting a causal role for *FBXW11* loss-of-function mutations in autoinflammatory disease.

**Funding:** This research was supported by grants from the European Union, ZonMW and the Radboud Institute for Molecular Life Sciences.

## Introduction

Although we inherit the vast majority of genomic variants from our parents, a small fraction of variants arises *de novo* during parental gametogenesis or after zygosis (1). The biological rate at which these variants develop in humans translates to an average of 50 to 100 *de novo* single nucleotide variants (SNVs) per genome per generation, usually only one or two of which affect coding regions (1, 2). *De novo* variants (DNVs) are often very rare or unique (absent from population databases) and have a higher *a priori* chance to be pathogenic than inherited variants (3, 4). In contrast to inherited variants, DNVs emerge between two generations and are subjected to limited evolutionary selection pressure that would normally purify damaging mutations (1). DNVs affecting nucleotides or genes that have been targeted by strong purifying selection can therefore be highly damaging to their respective non-redundant biological functions, as has been shown for genes involved in innate immunity, an ancient host defence mechanism that developed under constant environmental selection pressure by microorganisms (4, 5).

Therefore, DNVs are important candidates to pursue, particularly as a cause for rare sporadic diseases (2, 4, 6). The presence of such candidate DNVs can currently be assessed by trio-based sequencing, in which the patient is sequenced together with the (healthy) parents (1). Most experience with systematic assessment of DNVs has been gained in the field of developmental disorders, in which DNVs are evaluated diagnostically and have been shown to constitute up to 50% of disease-causing mutations (6–8). However, the contribution of DNVs in the pathogenesis of other disorders, such as inborn errors of immunity (IEI), is less clear.

DNVs as the underlying cause in IEI patients have been widely reported in literature, but most of these mutations have been predominantly determined to have originated *de novo* through subsequent segregation analysis and not by systematic trio-based sequencing (9–13). IEI can present at different stages of life with a variable phenotype ranging from recurrent, life-threatening infections to immune dysregulation and cancer (10, 14). Particularly in IEI patients with early-onset and severe complex phenotypes, there is an increased chance to identify an underlying causative DNV (4, 15). Moreover, post-zygotically or somatically arising DNVs are recognized as an important underlying cause for IEI patients with autoinflammatory disease (16–24). The potentially added value of systematic DNV assessment in suspected IEI patients is supported by the findings of an international cohort study, which reported that patient-parent trio sequencing resulted in a diagnosis in 44% of cases, compared to 36% by single whole exome sequencing (WES) (9). However, trio-based sequencing has not yet been implemented as part of the routine diagnostic procedure of IEI patients.

The current study has aimed to further explore the potential added value of systematic assessment of candidate DNVs in a retrospective cohort of 123 sporadic, suspected IEI patients that underwent trio-based WES.

## Methods

### Patients and samples

This retrospective cohort study was conducted on cases encountered in Genome Diagnostics between May 2013 and November 2021 at the Department of Human Genetics in the Radboud University Medical Center (RUMC). In some these cases, a patient-parent trio design was requested by the referring clinician. Patient-parent trios were selected for exome-wide *de novo* variant (DNV) analysis when fulfilling the following criteria: 1) the patient’s phenotype was sporadic, 2) the clinical description was suspect for an inborn error of immunity (IEI), and 3) the *in silico* IEI whole exome sequencing (WES) panel was requested and analysed. The *in silico* IEI gene panel of the RUMC is periodically updated after literature review and currently encompasses 456 genes (version DG3.1.0 (25)). During the study period, the *in silico* IEI WES panel was analysed in 146 patient-parent trios, of which 123 trios met the inclusion criteria (Figure 1).

**Figure 1.**
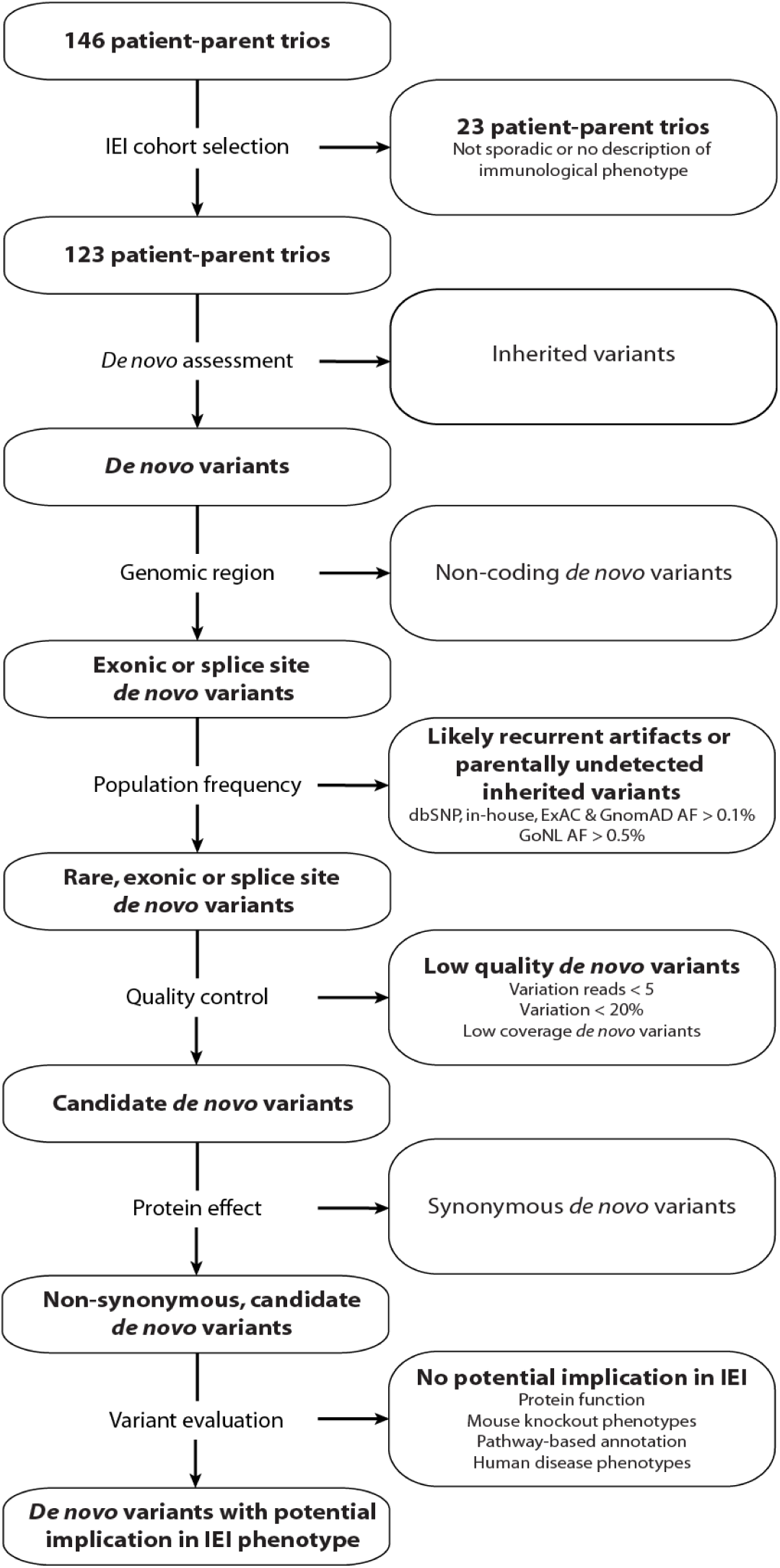
Schematic overview of patient inclusion, *de novo* variant filtering strategy and variant evaluation. Of the 146 eligible patient-parent trios, 123 trios met the inclusion criteria for this IEI cohort study. Whole exome sequencing data from these patient-parent trios was subjected to standardised variant filtering to retain candidate *de novo* variants. Subsequently, non-synonymous DNVs were systematically evaluated at variant and gene level for their potential involvement in the patient’s immunological phenotype. ***Figure 1 - table supplements 1-3, and figure supplement 1.*** Abbreviations: IEI = inborn errors of immunity dbSNP = Single Nucleotide Polymorphism Database. ExAC = Exome Aggregation Consortium. GnomAD = Genome Aggregation Database. AF = allele frequency. GoNL = Genome of the Netherlands.

As described previously (10), patients and their parents provided written informed consent for *in silico* IEI WES gene panel analysis with or without exome-wide variant analysis in line with the diagnostic procedure and clinical question, as approved by the local ethics committee (medisch-ethische toetsingscommissie Oost-Nederland). This research is in compliance with the principles of the Declaration of Helsinki (26).

For the exome-wide DNV analysis in this study, WES data of all subjects was pseudonymised. This entailed that patient DNA numbers were enciphered at random to ascending numbers for our study by a Genome Diagnostics member from the department of Human Genetics in the RUMC and clinical descriptions were condensed. Prior to this study, one patient was published as a clinical case report by D’hauw *et al.*, 19 patient-parent trios were included in the IEI cohort of Arts *et al.*, and one trio was part of a study by Konrad *et al.* (Figure 1 – table supplement 1) (10, 27, 28).

### Diagnostic whole exome sequencing

WES was performed as described previously with minor modifications (29). In brief, genomic DNA samples isolated from whole blood were processed at either the Beijing Genomics Institute (BGI) Europe (BGI Europe, Copenhagen, Denmark) or the in-house sequencing facility. All samples were enriched for exonic DNA using Agilent (Agilent Technologies, Santa Clara, CA, United States) or Twist (Twist Bioscience, San Francisco, CA, United States) exome kits. DNA samples at BGI were sequenced on Illumina HiSeq4000 (Illumina Sequencing, San Diego, CA, United States) or DNBseq (MGI Tech, Shenzhen, China). In-house DNA samples were sequenced on Illumina NovaSeq6000 (Illumina Sequencing). Sequencing was performed with 2×100 base pair (DNBseq) or 2×150 base pair (HiSeq4000 and NovaSeq6000) paired-end sequencing reads. The average median sequence coverage was 124x with an average of 96% target coverage greater than 20x (Figure 1 – table supplement 1).

Downstream processing was performed by an automated data analysis pipeline, including mapping of sequencing reads to the GRCh37/hg19 reference genome with the Burrows-Wheeler Aligner algorithm and Genome Analysis Toolkit variant calling and additional custom-made annotation (30, 31). The DeNovoCheck tool is part of the custom-made annotation and was used to align variants called in each member of the patient-parent trios, providing an indication whether variants were inherited or *de novo* (29). Subsequently, all single nucleotide variants (SNVs) or small insertion-deletions (indels) were annotated by a custom, in-house annotation pipeline. Copy number variants (CNVs) were assessed by the copy number inference from exome reads (CoNIFER) method, as of 2018. (32).

Subsequently, variants in genes included in the *in silico* IEI panel were filtered to retain both inherited and *de novo* coding, non-synonymous variants with population frequencies below 1% in our in-house database or population databases (GnomAD and dbSNP) (33, 34). Variant prioritisation was performed by clinical laboratory geneticists of the Department of Human Genetics at the RUMC. SNVs, small indels or CNVs that may (partially) be related to the phenotype were classified (five-tier classification) and reported according to guidelines of the Association for Clinical Genetic Science and the American College of Medical Genetics and Genomics (ACMG) (35, 36) (Table 2A). Variants that were denoted or classified as carriership of a variant in a known recessive disease gene, known risk factors or variants of uncertain significance or (likely) pathogenic variants in disease genes other than those associated with IEI, and candidate variants identified after exome-wide variant analysis were additionally reported and are listed in Table 1A – table supplement 1.

### *De novo* variant analysis

As part of this study, an additional exome-wide re-analysis directed towards the identification of DNVs in 123 patient-parent trios was performed. For this, a standardised variant filtering strategy was applied using R Studio version 3.6.2. Variants were filtered to retain rare (<0.1% allele frequency in our in-house database and the population databases from Exome Aggregation Consortium (ExAC), Genome Aggregation Database (GnomAD) and dbSNP as well as <0.5% in the Genome of the Netherlands (GoNL) database), coding, non-synonymous possible DNVs, as annotated by the DeNovoCheck tool (Figure 1) (29, 33, 34, 37, 38). Quality control steps excluded variants with <5 variation reads, <20% variant allele frequency or low coverage DNVs. Subsequently, synonymous SNVs and small indels were excluded from the analysis. The remaining DNVs were considered candidate DNVs and are listed in Figure 1 – table supplement 2. These candidate DNVs were prioritised and systematically evaluated using variant and gene level metrics, containing database allele frequencies (including DNV counts in other datasets via denovo-db), nucleotide conservation, pathogenicity prediction scores, functional information and possible involvement in the immune system based on mouse knockout models, pathway-based annotation (i.e., Gene Ontology terms), and literature studies (33, 39–41). In addition, splice site DNVs were analysed using the Alamut Visual Software version 2.13 (SOPHiA GENETICS, Saint Sulpice, Switzerland), which provides splicing prediction tools including SpliceSiteFinder-like, MaxEntScan, NNSPLICE, GeneSplicer and ESE tools.

### FBXW11 functional validation experiments

#### Epstein–Barr virus (EBV)-B cell lines

Venous blood was drawn from patient 53 and collected in lithium heparin tubes. Epstein–Barr virus (EBV)-transformed B cell lines were created following established procedures (42). EBV-transformed lymphoblastoid cell lines (EBV-LCLs) from the patient and a healthy control were grown at 37°C and 7.5% CO_2_ in RPMI 1640 medium (Dutch Modification, Gibco; Thermo Fisher Scientific, Inc., Waltham, MA, United States) containing 15% foetal calf serum (FCS; Sigma-Aldrich, St Louis, MO, United States), 1% 10.000U/μl penicillin and 10.000 μg/μl streptomycin (Sigma-Aldrich), and 2% HEPES (Sigma-Aldrich). The EBV-LCLs were cultured at a concentration of 10×10^6^ in 150cm^2^ culture flasks (Corning, Corning, NY, United States) and treated with or without cycloheximide at 0.1% (20mL/20mL medium; Sigma-Aldrich) for four hours. Cell pellets were then spun down, washed with PBS, snap-frozen in liquid nitrogen and stored at -80°C.

#### RNA splicing effect

RNA was isolated from the EBV-B cell pellets using the RNeasy Mini isolation kit (Qiagen, Hilden, Germany) according the manufacturer’s instructions. All obtained RNA was used for cDNA synthesis with the iScript™ cDNA Synthesis Kit (Bio-Rad, Hercules, CA, United States). A primer set was designed (Primer3web, version 4.1.0) to span exon 11 to 13 of *FBXW11*, with the following sequences: Forward 5’-GAGAGCCGGAATCAGAGGTG-3’; Reverse 5’-GAATTGGTCCGATGCATCCG-3’. Subsequently, RT-PCR was performed using the AmpliTaq Gold™ 360 Master Mix (Life Technologies, Carlsbad, CA, United States). The amplified PCR products and Orange G ladder were electrophoresed on a 2% agarose gel with GelRed, and the resulting bands were cut out and analysed with Sanger sequencing.

#### Ex vivo peripheral mononuclear blood cell (PBMC) experiments

Venous blood was drawn and collected in EDTA tubes. Immune cell isolation was conducted as described elsewhere (43). In brief, PBMCs were obtained from blood by differential density centrifugation, diluted 1:1 in pyrogen-free saline over Cytiva Ficoll-Paque Plus (Sigma-Aldrich). Cells were washed twice in saline and suspended in cell culture medium (Roswell Park Memorial Institute (RPMI) 1640, Gibco) supplemented with gentamicin, 10 mg/mL; L-glutamine, 10 mM; and pyruvate, 10mM. *Ex vivo* PBMC stimulations were performed with 5×10^5^ cells/well in round-bottom 96-well plates (Greiner Bio-One, Kremsmünster, Austria) for 24 hours in the presence of 10% human pool serum at 37°C and 5% carbon dioxide. For cytokine production measurements, cells were treated with *Candida albicans* yeast (1×10^6^/mL), lipopolysaccharide (LPS, 10 ng/mL), *Staphylococcus aureus* (heat-killed, 1×10^6^/mL) or TLR3 ligand Poly I:C (10 μg/mL) or left untreated in regular RPMI medium. After the incubation period and centrifugation, supernatants were collected and stored at −20°C until the measurement using enzyme-linked immunosorbent assay (ELISA).

For flow cytometry experiments, PBMCs were cultured in U-bottom plates at a final concentration of 1×10^6^ cells in 200μL per well containing culture medium supplemented with 5% FCS (Sigma-Aldrich) at 37°C and 5% carbon dioxide. Subsequently, cells were stimulated with phorbol 12-myristate 13-acetate (PMA, 12.5 ng/mL, Sigma-Aldrich) and ionomycin (500 ng/mL, Sigma-Aldrich) in duplicate for 30 minutes.

#### Flow cytometry

PBMC suspensions were transferred to a V-bottom plate while pooling the duplicates. Following centrifugation for 2.5 minutes, cell surface markers were stained in the dark for 30 minutes at 4°C with a monoclonal antibody mix containing anti-CD3-ECD (1:25; Beckman Coulter, Brea, CA, United States), anti-CD4-BV510 (1:50; BD Bioscience, Franklin Lakes, NJ, United States), anti-CD8-APC Alexa Fluor™ 700 (1:400; Beckman Coulter), and anti-CD14-FITC (1:50; Dako; Agilent Technologies). Subsequently, cells were washed twice with flow cytometry buffer (FCM buffer, 0.2% BSA in PBS) and fixed (BD Biosciences Cytofix, 554655) for 10 minutes at 37°C. Next, cells were washed and permeabilised with perm buffer IV (1:10 diluted with PBS, BD Biosciences Phosflow, 560746) for 20 minutes on ice in the dark. Cells were then stained intracellularly with anti-NF-κB p65 (pS529)-PE antibody (1:50; eBioscience; Thermo Fisher Scientific, Inc., Waltham, MA, United States) for 20 minutes at 4°C. After washing the cells twice in FCM-buffer, the suspensions were measured on a Beckman Coulter Navios EX Flow Cytometer using Navios System Software. Cell immunophenotypes were analysed using Kaluza Analysis Software version 2.1 (Beckman Coulter). The mean fluorescent intensities (MFIs) were calculated using the median pNF-κB p65 expression levels within the gated immune cell populations of interest.

#### Cytokine measurements

Levels of cytokines IL-1β, IL-6 and TNFα were determined in supernatants of stimulated PBMC cultures according to the instructions of the manufacturer (Duoset ELISA; R&D Systems, Minneapolis, MN, United States).

## Results

### Cohort characteristics

This retrospective cohort study systematically re-analysed patient-parent trio whole exome sequencing (WES) data of 123 patients with suspected inborn errors of immunity (IEI) with the aim to identify (likely) pathogenic *de novo* single nucleotide variants (SNVs) or small insertion-deletions (indels) (Figure 1). Included IEI patients had a median age of 9 years (IQR 2-17) and two-thirds of the cases were below 18 years of age (Table 1A). The sex distribution among patients was roughly equal. Classification of IEI phenotypes according to the International Union of Immunological Societies (IUIS) indicated that most cases presented with autoinflammatory syndromes, followed by immune dysregulation and combined, predominantly syndromal immunodeficiencies (14). Eight patients remained unclassified due to limited clinical data.

**Table 1A.**
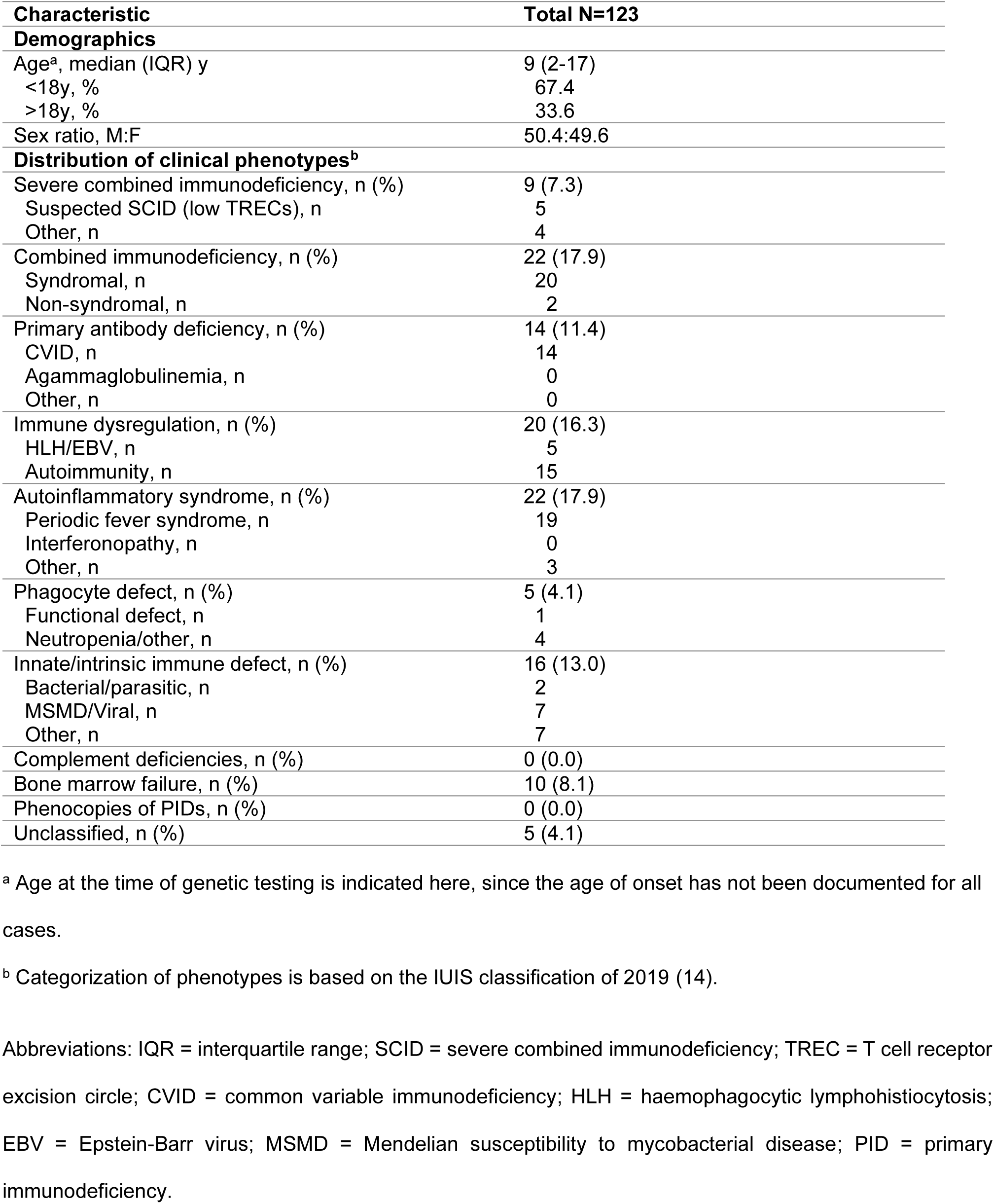
Patient cohort characteristics. Demographic and phenotypic characteristics of the 123 patients included in this cohort of inborn errors of immunity.

### Genetic variants reported after routine diagnostic whole exome sequencing analysis

Following routine diagnostic WES, potential disease-causing SNVs and/or copy number variants (CNVs) were reported in 36 index patients (Table 1B). Twenty-four patients were carriers of recessive disease alleles, previously characterised risk factors, variants of uncertain significance (VUS) or (likely) pathogenic variants affecting established disease genes other than those associated with IEI (Table 1B). Of note, three of these patients carried *de novo* CNVs (patient 21, 69 and 115).

**Table 1B.** Genetic findings after routine diagnostic panel analysis. Genetic variants reported after routine diagnostic whole exome sequencing analysis of the 123 patients included in this cohort of inborn errors of immunity. ***Table 1B – table supplement 2*.**

|  |  |
| --- | --- |
| **Total cases in which a genetic variant was reported, n (%)** | **36 (29.3)** |
| **Likely pathogenic mutation, n (%)** | **19 (15.4)** |
| Within IEI gene panel, n (%) | 12 (9.8) |
| Beyond IEI gene panel, n (%) | 7 (5.7) |
| **Other variants, n (%)** | **17 (13.8)** |
| Risk factor, n (%) | 6 (4.9) |
| Carriership recessive allele, n (%) | 7 (5.7) |
| Variant of unknown significance, n (%) | 9 (7.3) |

In 12 patients, (likely) pathogenic SNVs were identified in known IEI genes that (partially) explain the patient’s immunological phenotype (Table 1B, details shown in Table 2A). While the majority of variants was inherited, one patient with Muckle-Wells syndrome (patient 59) carried a *de novo* missense variant in *NLRP3* (NM_001079821.2:c.1049C>T p.(Thr350Met)). This variant has previously been described in patients with Muckle-Wells syndrome (44, 45). Consequently, the *NLRP3 de novo* variant (DNV) was classified as pathogenic (35, 36).

**Table 2A.**
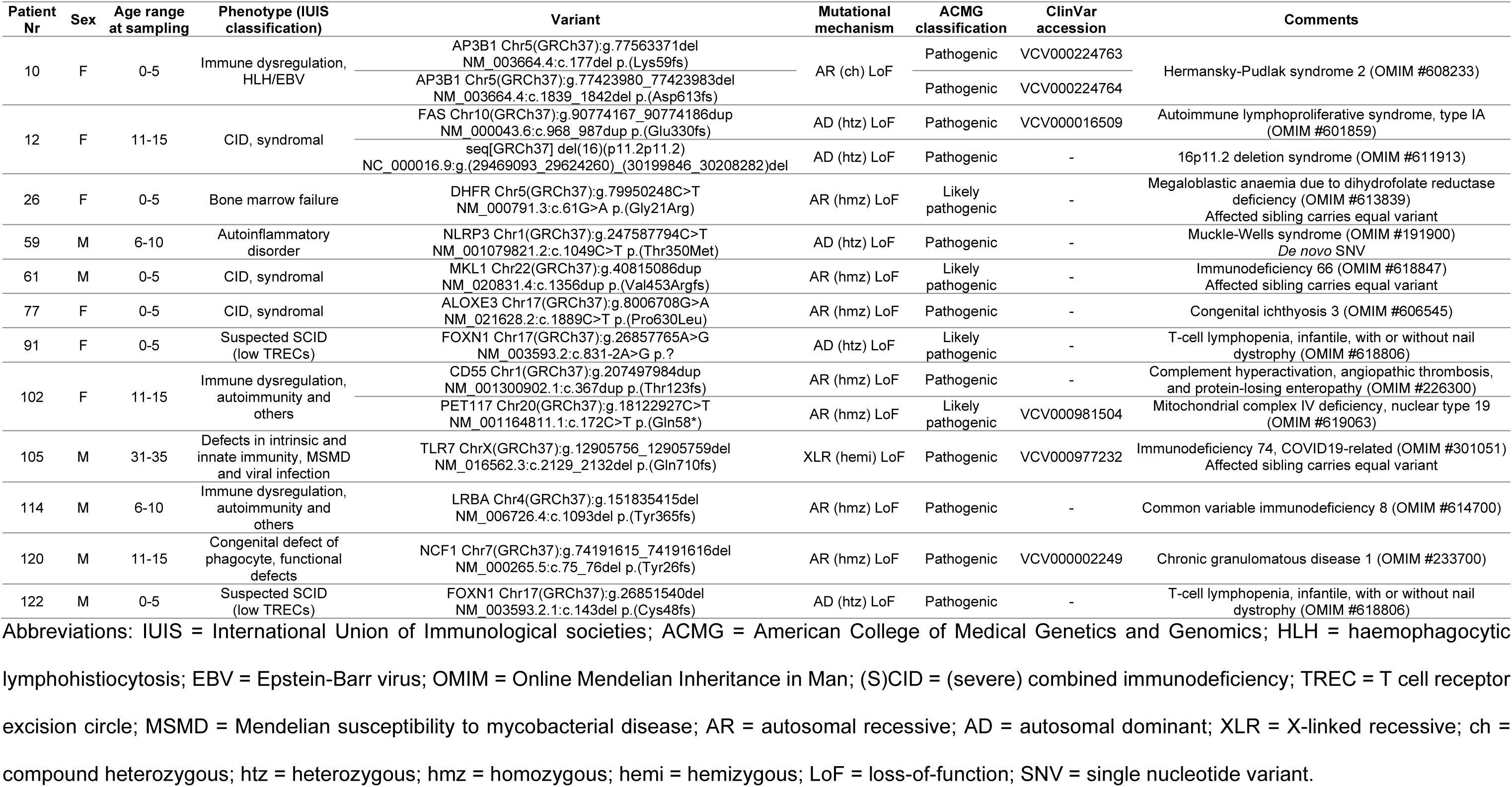
Patients with previously reported single nucleotide variants, small insertion-deletions, or copy number variants that may (partially) explain the patient’s immunological phenotype. Listed variants were identified prior to this cohort study in the scope of routine diagnostic inborn errors of immunity gene panel analysis of each patient included in this cohort.

Overall, routine diagnostic WES analysis provided a likely molecular diagnosis for (part) of the phenotype in 19 patients (15.4%) (Table 1B, Table 2A).

### Rare, non-synonymous *de novo* variants in novel IEI candidate genes

Next, exome-wide re-analysis was performed on WES data of all 123 sporadic IEI cases and their parents to systematically identify and interpret DNVs in novel IEI genes. Automated DNV filtering retained a total of 187 candidate DNVs that were rare and located in either exonic or splice site regions (the complete list can be found in Figure 1 – table supplement 2). The total number of candidate DNVs among patients ranged between zero and six (Figure 1 – figure supplement 1). Moreover, the average number of candidate DNVs was comparable to recent literature (Figure 1 – table supplement 3). Of these candidate DNVs, 136 were non-synonymous and therefore more likely to exert an effect on protein function (Figure 1 – figure supplement 1). Two pairs of patients carried candidate DNVs in the same gene, *GIGYF1* (patients 49 and 83) and *MAP3K10* (patients 98 and 118). However, these patients did not share phenotypic features and the function of the proteins encoded by these genes could not be linked to the respective patient phenotype.

Subsequently, all non-synonymous candidate DNVs were systematically evaluated based on information on variant and gene level, leading to the selection of 14 candidate DNVs potentially causing IEI (Table 2A and B), including the above-mentioned variant in the known IEI gene *NLRP3*. The 13 novel IEI candidate DNVs were found in patients with different IEI phenotypes, although three subtypes reoccurred: predominantly antibody deficiency (hypogammaglobulinemia), autoinflammatory disorder and bone marrow failure. Candidate DNVs that were considered most promising based on variant and gene level metrics are presented in more detail in the following paragraphs.

**Table 2B.**
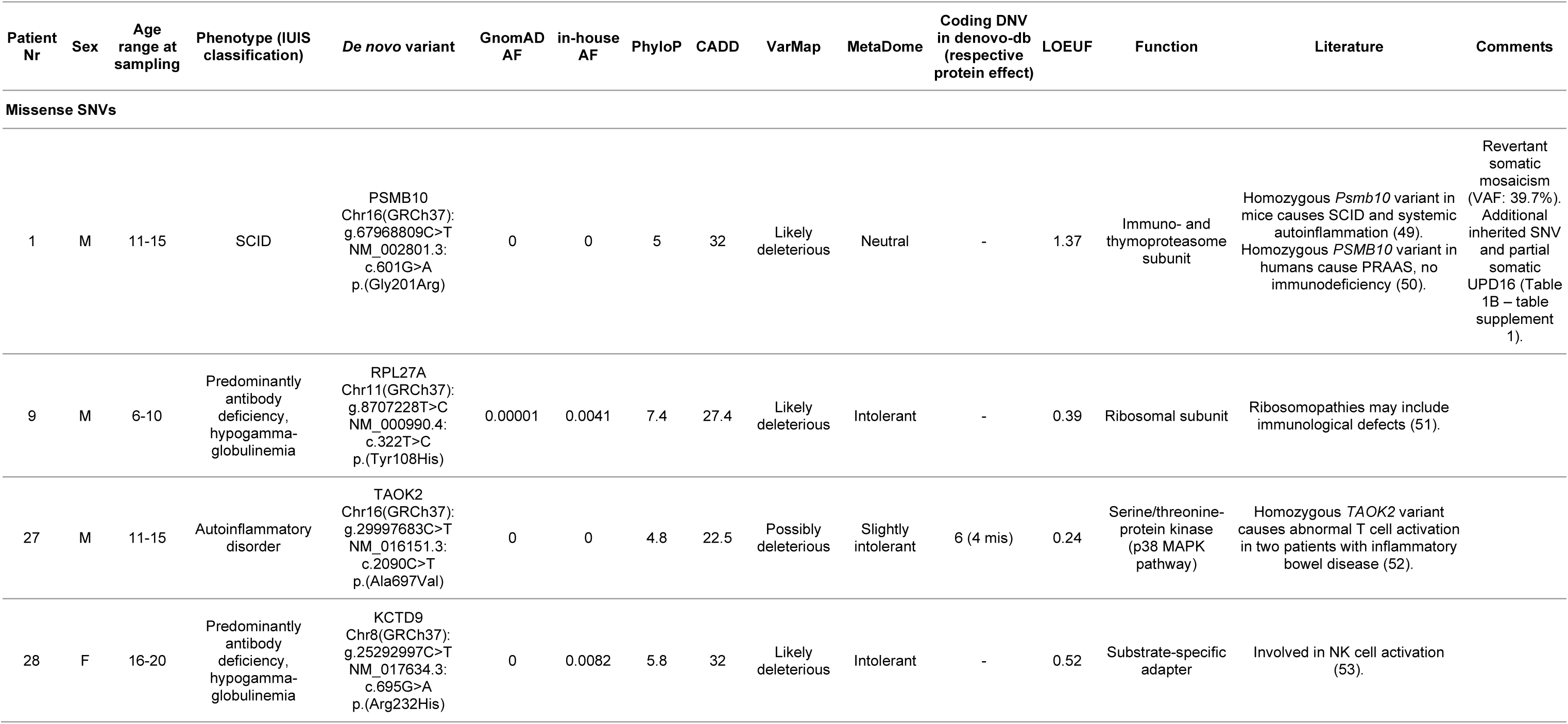
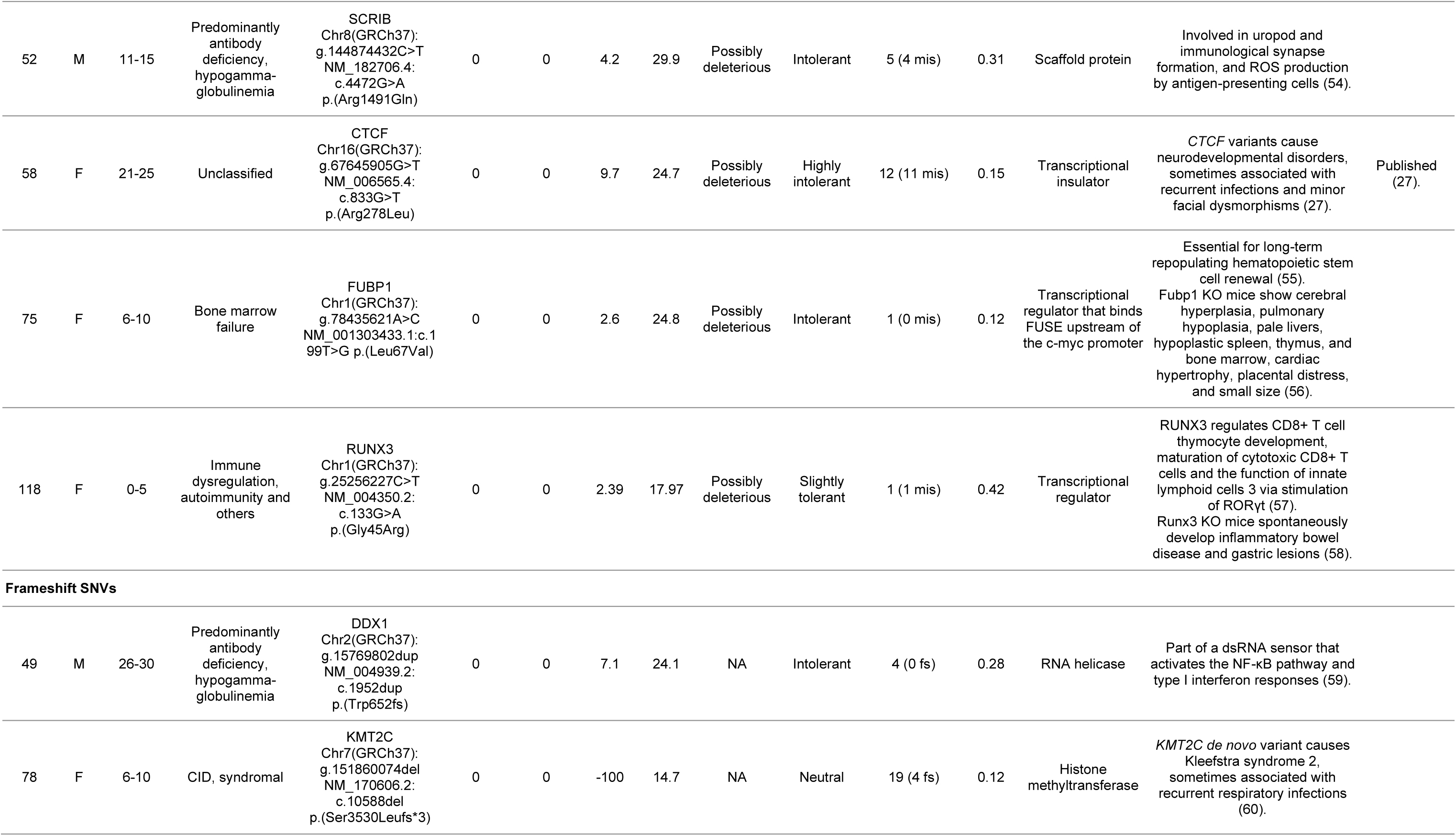
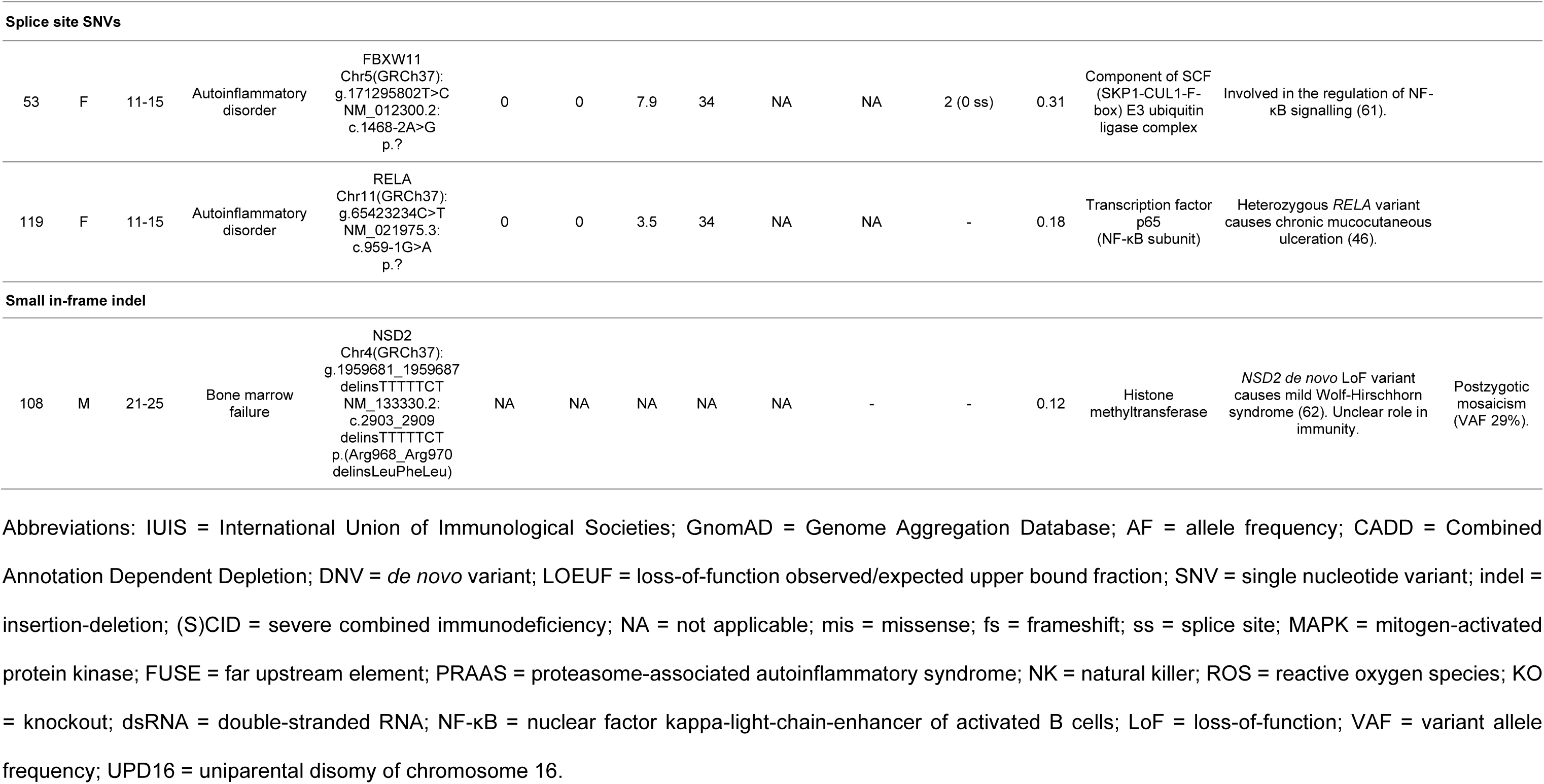
Identification of 13 heterozygous, rare and non-synonymous candidate *de novo* variants with immunological implication. The 136 non-synonymous candidate *de novo* variants were systematically evaluated based on the potential to be damaging to gene function and the involvement in the patient’s immunological defect. For this, variant and gene level metrics, containing database allele frequencies (including denovo-db), nucleotide conservation, pathogenicity prediction scores, functional information and possible involvement in the immune system based on mouse knockout models, pathway-based annotation (i.e., Gene Ontology terms) and literature studies were summarised (33, 39–41).

In a patient with an autoinflammatory phenotype characterised by mucocutaneous ulceration of mouth and genital area (patient 119, Table 2B), a DNV was located in the canonical splice acceptor site preceding exon 10 of *RELA* (NM_021975.3:c.959-1G>A). The guanine to adenine change was predicted to compromise the splice acceptor site by transferring it to the first guanine of exon 10, leading to an out-of-frame exon. The resulting frameshift was therefore assumed to cause a reduction in functional RelA protein by nonsense-mediated decay. RelA is also known as p65 and is critically involved in nuclear factor kappa-light-chain-enhancer of activated B cells (NF-κB) heterodimer formation and consequent activation of NF-κB-mediated proinflammatory signalling. RelA haploinsufficiency has been reported as a cause of chronic mucocutaneous ulceration and familial Behçet’s disease (46, 47). Badran *et al.* reported a family of four affected family members with mucocutaneous ulceration harbouring a mutation in the canonical donor splice site of exon 6 (NM_021975:c.559+1G>A), likely leading to a premature stop codon and haploinsufficiency (46). Both, the phenotype and proposed mutational mechanism in our patient, match this description. Although *RELA* has already been reported as an IEI gene in a previous IUIS classification (48), it was not yet listed in the IEI *in silico* gene panel of our Department of Human Genetics (25), because evidence was considered insufficient at the time. Based on these arguments, this DNV has only now been classified as pathogenic, which could carry implications for therapy with anti-tumour necrosis factor alpha (TNFα) inhibitors (47).

In addition, a private *de novo* missense variant in *PSMB10* was observed in a patient with clinically diagnosed Omenn syndrome with severe combined immunodeficiency (SCID), ectodermal dysplasia, alopecia, hypodontia and anonychia (patient 1, Table 2B). The clinical phenotype of this patient has been previously reported (28). The DNV was predicted to be pathogenic based on the majority of variant and gene level metrics. In additional data that was available from a previously performed single nucleotide polymorphism (SNP) micro-array, it was shown that the genomic location of *PSMB10* was spanned by a partial somatic uniparental disomy of chromosome 16 (UPD16) (manuscript in preparation). *PSMB10* encodes the β2i-subunit of the immuno- and thymoproteasome, and mutations leading to a loss of PSMB10 protein function have been associated with severe immunological defects (49). A homozygous *PSMB10* variant (p.Gly170Trp) has been shown to induce SCID and systemic autoinflammation in mice, while heterozygous mice only had a T cell defect (49). In addition, a 3-year-old Algerian female with autoinflammatory signs suggestive of proteasome-associated autoinflammatory syndrome (PRAAS) was shown to harbour a homozygous missense *PSMB10* variant (NM_002801.3:c.41T>C p.(Phe14Ser)), leading to disturbed formation of the 20S proteasome (50). Mutations in other genes encoding B subunits of the immunoproteasome, including *PSMB8* and *PSMB9*, are known autosomal recessive causes of PRAAS (50, 63, 64). Furthermore, it is interesting to consider that the amino acid position of our variant is located in proximity to the position of the p.(Gly170Trp) variant in the mouse orthologue (49). However, the functional consequence and pathogenic relevance of the candidate DNV in *PSMB10* remain unknown; it is possible that the DNV acts through a novel autosomal dominant mutational mechanism.

Furthermore, in a patient with common variable immunodeficiency (CVID) due to a B cell maturation defect, auto-immune cytopenia, polyclonal T cell large granular lymphocytes in the bone marrow, recurrent viral infections, psoriasis and alopecia areata, another candidate DNV was identified in *DDX1* (patient 49, Table 2B). This frameshift variant was predicted to cause loss of DDX1 protein function. *DDX1* encodes an RNA helicase, which is part of a double-stranded RNA sensor that activates the NF-κB pathway and type I interferon responses (59). Moreover, DDX1 is involved in the regulation of hematopoietic stem and progenitor cell homeostasis (59, 65).

Another frameshift DNV was carried by a patient with a syndromal combined immunodeficiency characterised by recurrent ear infections, developmental delay, low-average intelligence level and facial dysmorphism (patient 78, Table 2B). The variant was predicted to lead to a loss-of-function (LoF) of the KMT2C protein, which acts as a histone methyltransferase. In humans, *de novo* LoF mutations in KMT2C are associated with Kleefstra syndrome 2, a neurodevelopmental disorder (60). Two of the six individuals described in that study were reported to have recurrent respiratory infections (60). The occurrence of immunological symptoms in patients with mutations in chromatin-regulating genes is increasingly being recognized in the field of intellectual disability (ID) (66, 67). Therefore, more in-depth characterisation of patients with *KMT2C* mutations and predominant ID phenotypes might indicate (mild) immunological phenotypes that overlap with the phenotype of our patient, in support of pathogenicity of the observed DNV.

Lastly, a DNV was identified in *FBXW11,* carried by a patient with an autoinflammatory disorder characterised by recurrent periodic fever and severe headaches (patient 53, Table 2B). *FBXW11* encodes a component of SCF (SKP1-CUL1-F-box) E3 ubiquitin ligase complex, TrCP2, that is involved in the regulation of NF-κB signalling through the ubiquitination of several of its components (61, 68). An important function of both the TrCP1 and TrCP2 isoforms is the regulation of IκBα degradation, leading to subsequent activation of NF-κB and release of pro-inflammatory cytokines (69, 70). The identified DNV affected the canonical splice acceptor site preceding exon 12 (NM_012300.2:c.1468-2A>G). This DNV was predicted to lead to skipping of exon 12 based on splicing prediction by the Alamut Visual Software and to be deleterious by all utilised *in silico* prediction tools. The predicted RNA splicing defect leading to an in-frame, shortened RNA transcript was confirmed in Epstein-Barr virus (EBV) transformed B cells from the patient (Figure 2).

**Figure 2.**
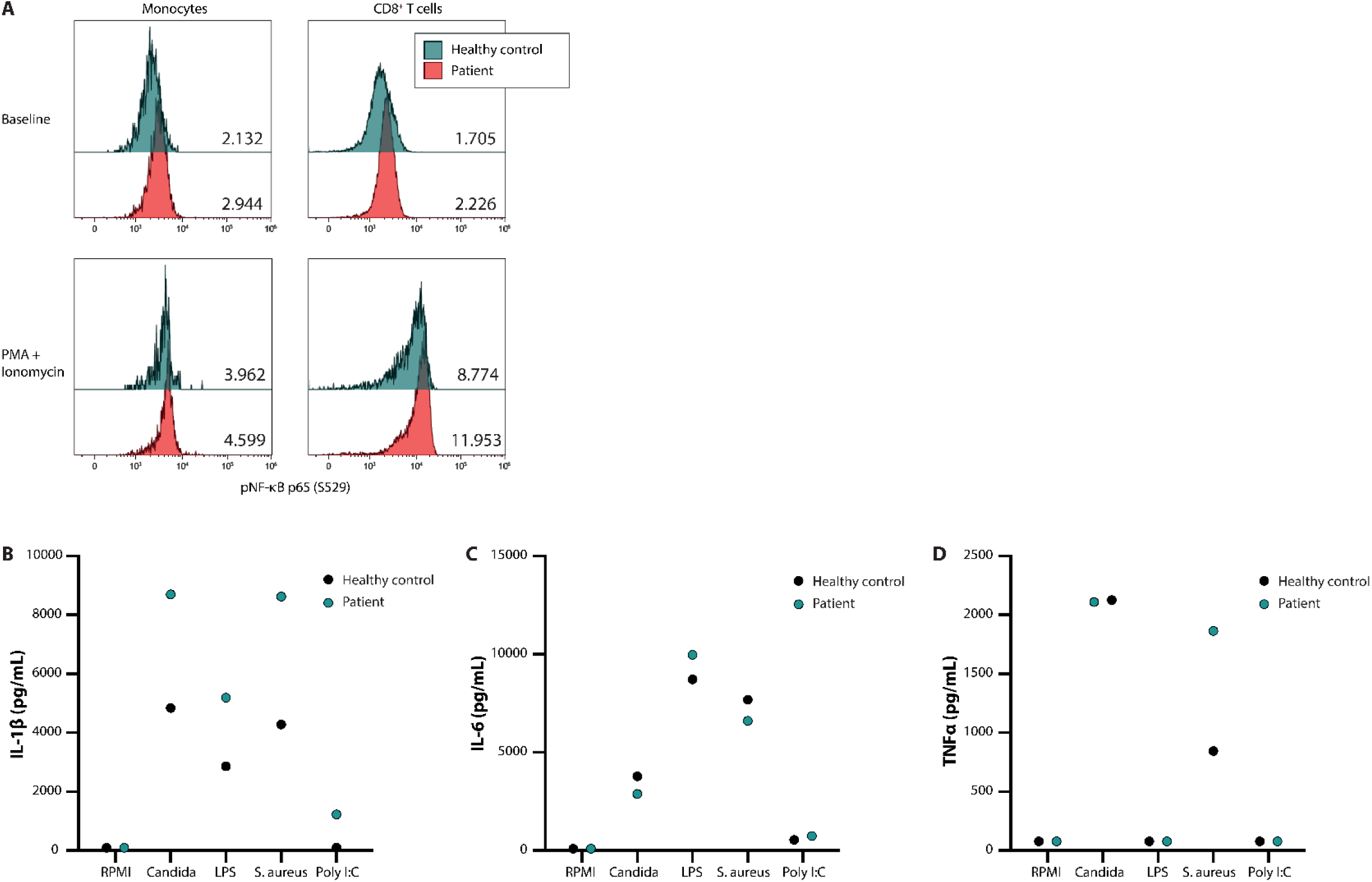
NF-κB signalling and production of innate cytokines upon *ex vivo* PBMC stimulation. Panel A shows the median fluorescence intensity expression levels of pNF-κB p65 (S529) in peripheral blood CD14+ monocytes and CD8+ T cells from a healthy control (blue) and patient 53 (red), in the absence (baseline) or presence of phorbol 12-myristate 13-acetate and ionomycin stimulation, with the absolute values indicated in the lower right corner. Panel B, C and D display the production of IL-1β, IL-6 and TNFα, respectively, after *ex vivo* stimulation for 24 hours. ***Figure 2 – figure supplement 1; Figure 2B-D – source data 1.***

The other candidate DNVs will not be described in detail here, as there is insufficient evidence to suggest pathogenicity or a genotype-phenotype relationship. Future discovery of cases with DNVs in the presented genes and overlapping clinical phenotypes could encourage further in-depth research into the possible mutational mechanisms.

### Functional validation of *FBXW11 de novo* variant

In addition to systematic DNV analysis, we have selected the candidate DNV in *FBXW11* for functional validation as part of this study to provide further evidence for a causal genotype-phenotype relationship (patient 53, Table 2B). As such, the putative effects on NF-κB signalling and the downstream production of pro-inflammatory cytokines were investigated in peripheral blood mononuclear cells (PBMC) extracted from the patient and a healthy control. In unstimulated PBMC of the patient showed higher levels of phosphorylated NF-kB p65 compared to the control. *Ex vivo* stimulation of these PBMC with phorbol 12-myristate 13-acetate (PMA) and ionomycin for 30 minutes led to higher NF-κB activation, reflected by p65 phosphorylation fluorescence intensity measured by flow cytometry, as compared to the healthy control (Figure 2, panel A). The greatest difference was observed in the lymphocyte subset, particularly in CD8+ T cells (Figure 2, panel A). Subsequently, the downstream production of the cytokines IL-1β, IL-6 and TNFα was investigated. The patient-derived PBMC produced more IL-1β upon *in vitro* stimulation with the heat-killed pathogens *Candida albicans* and *Staphylococcus aureus*, the TLR4 agonist lipopolysaccharides (LPS) and the TLR3 ligand Poly I:C after 24 hours, as compared to the healthy control (Figure 2, panel B). This trend was not observed for the production of IL-6 and TNFα (Figure 2, panel C and D). These results indicate that the *FBXW11* DNV leads to a splicing defect with skipping of exon 12, resulting in a shorter transcript and increased NF-κB signalling and downstream IL-1β production.

## Discussion

We investigated the potential benefit of exome-wide trio-based sequencing over routine single whole exome sequencing (WES) analysis in a retrospective cohort of 123 patients with sporadic, suspected inborn errors of immunity (IEI). Systematic analysis of *de novo* single nucleotide variants (SNVs) and small insertion-deletions (indels) led to the identification of 14 candidate *de novo* variants (DNVs), of which two were in known IEI genes and classified as pathogenic (*NLRP3, RELA*). Of the 12 variants in potentially novel candidate genes for IEI, four were considered to be most likely pathogenic (*PSMB10, DDX1, KMT2C, FBXW11*) based on gene and variant level metrics. Additionally, we have provided functional evidence that the *FBXW11* splice site DNV led to skipping of exon 12 resulting in the transcription of an altered protein product and subsequent downstream activation of NF-κB signalling with higher IL-1β production capacity.

We have performed a systematic, exome-wide DNV analysis in selected patients with sporadic, suspected IEI. On average, these patients carried 0.89 non-synonymous DNVs in coding regions, a rate comparable to other, larger studies, indicating that an enrichment or depletion of DNVs in IEI patients is unlikely (8). Based on gene and variant level information, 14 DNVs (11.4%) were considered potential disease-causing candidates. This would result in a maximum solve rate of 23.6%, when combining the candidate DNVs with the 12 (likely) pathogenic inherited variants in known IEI genes and the three reported *de novo* copy number variants (CNVs) with unknown pathogenicity. Six of the candidate DNVs (4.9%) were considered likely or possibly pathogenic variants, while the consequence of the other nine DNVs (6.5%) was uncertain. Two DNVs were in IEI genes (*NLRP3, RELA*) listed in the most recent IUIS classification and were classified as pathogenic (35, 36, 48). The heterozygous *NLRP3* variant in patient 59 (p.Thr350Met) with Muckle-Wells syndrome had been reported in patients with an equal phenotype (44, 45). Similarly, the canonical splice site DNV affecting *RELA* in patient 119 with mucocutaneous ulceration was predicted to lead to a loss of the splice acceptor site and a subsequent frameshift, analogous to the previously demonstrated mutational mechanism for a canonical donor splice site variant that led to RELA haploinsufficiency causing the same phenotype (46).

Moreover, DNVs in the potentially novel IEI genes *PSMB10*, *DDX1*, *KMT2C* and *FBXW11* were considered the most promising candidate DNVs based on the predicted variant effect and immunological function of the respective gene. The private missense DNV in *PSMB10* was found in a patient with clinically diagnosed Omenn syndrome, showing phenotypic resemblance with features reported in a mouse model that investigated the effect of a mutation proximal to that of the patient in the human homologue (49). The presumed deleterious effect was further supported by the extremely rare occurrence of revertant mosaicism in this patient (unpublished data), *i.e.,* somatic and recurrent uniparental disomy 16q overlapping the *PSMB10* locus, suggesting a strong (cellular) effect of this variant. In addition, a *de novo* frameshift variant in the highly intolerant *DDX1* (pLI 0.994) was identified in a patient with hypogammaglobulinemia, hematopoietic cell lineage abnormalities and recurrent infections. Although a causal genotype-phenotype relationship remains unclear, it has been reported that DDX1 plays a role in NF-κB signalling, type I interferon responses, and the regulation of hematopoietic stem and progenitor cell homeostasis (59, 65). Furthermore, the *de novo* frameshift variant in *KMT2C* was detected in a patient with combined immunodeficiency and a neurodevelopmental phenotype, displaying partial phenotypic overlap with Kleefstra syndrome type 2 that has already been associated with *de novo* mutations in *KMT2C* (60).

Another promising candidate DNV in a potentially novel IEI gene was identified in a patient with periodic fever and was located in the highly conserved *FBXW11* (pLI 0.976). This DNV affected the canonical splice acceptor site preceding exon 12 and was shown to create a splice defect leading to exon skipping with a shortened transcript that retained expression at the RNA level. Exon 12 encodes a component of the WD40 repeat domain, which is involved in substrate recognition (71). *De novo* missense and nonsense variants in *FBXW11* have been previously described in patients with a neurodevelopmental syndrome with abnormalities of the digits, jaw and eyes (72). These variants have been shown to compromise substrate recognition or binding of the Wnt and Hedgehog signalling developmental pathways. In our patient with a distinct autoinflammatory phenotype, we hypothesised a specific functional effect on NF-κB signalling. *FBXW11* encodes β-TrCP2, a component of the SCF (SKP1-CUL1-F-box) E3 ubiquitin ligase complex that mediates the ubiquitination of IκBα and consequently stimulates canonical NF-κB signalling (73). It was previously shown that the abundance of β-TrCP, which includes the highly homologous isoform β-TrCP1, affects the steady-state concentration of NF-κB and its dynamics on stimulation (73). In peripheral blood mononuclear cells (PBMC) extracted from the patient, we demonstrated that the phosphorylation of the NF-κB subunit p65 was constitutively higher in monocytes and CD8+ T cells as compared to a healthy control, which suggests a functional effect of the *FBXW11* variant. This effect is further substantiated by the observation of increased p65 phosphorylation and downstream production of IL-1β after stimulation with pathogens and a TLR3 ligand in the patient. However, a note of caution should be made regarding n=1 studies, as we cannot exclude that the difference is due to normal inter-individual biological variability. These results suggest that NF-κB signalling was aberrantly increased in the patient, a mechanism that has been shown to be involved in the pathogenesis of other monogenic autoinflammatory disorders known as relopathies (74). Although it is likely that the *de novo* variant in *FBXW11* plays a role in the immunological phenotype, further experiments addressing the effect of this DNV on IκBα degradation, substrate recognition and TrCP protein abundance should be undertaken to provide conclusive evidence.

To our knowledge, two other cohort studies have systematically performed trio-based sequencing in IEI patients as part of their study design, although patients were not pre-selected based on sporadic phenotypes (9, 75). Stray-Pedersen *et al.* conducted a large international cohort study to investigate the usefulness of WES in IEI patients from 278 families, which included 39 patient-parent trios (9). The authors reported a molecular diagnosis in 40% of the patients, including 15 (13.6%) *de novo* mutations, of which 4 were identified by trio-based analysis and 11 after segregation analysis. The additional value of trio-based sequencing is indicated by the higher detection rate compared to that of the single cases followed by segregation analysis of candidate variants (44 versus 36%), as well as the discovery of potentially novel IEI genes or expansion of the immunological phenotype. Furthermore, Simon *et al.* performed WES in a cohort of 106 IEI patients with a consanguineous background, including 26 patient-parent trios (75). A molecular diagnosis was established in 70% of the patients, including 13 (17.6%) *de novo* mutations, although it is unclear whether these variants were identified through trio-based sequencing or the segregation analysis that was performed for each variant. The authors conclude that trio-based sequencing does not lead to additional diagnostic benefit, although it should be argued that this is also not expected in a cohort of predominantly consanguineous patients (62.2%) with a higher *a priori* chance of autosomal recessive disease.

Multiple studies have highlighted the potential benefits of routine trio-based sequencing in IEI patients over single WES (3, 10, 15, 76). These advantages apply to patients with sporadic, severe phenotypes in particular, as has been shown for other rare diseases such as neurodevelopmental disorders (8). Trio-based sequencing constitutes an unbiased way to identify rare, coding DNVs that are by definition strong candidate variants. It could therefore improve candidate variant prioritisation both during *in silico* gene panel analysis as part of routine diagnostics, as well as during exome-wide analysis in a research setting. Furthermore, targeted DNV analysis could improve the detection of somatic variants, which is especially relevant in the field of monogenic autoinflammatory disorders (23). Somatic variants can be successfully identified by trio-based WES (77). However, this specific DNV subtype can be missed during routine analysis especially if the variant allele frequency (VAF) is below the set threshold during standard variant filtering, which is not required to filter out false-positive variants for a condensed set of potential DNVs. Another advantage of trio-based sequencing is that it provides direct segregation of inherited variants and enables determination of autosomal recessive compound heterozygosity or X-linked recessive disease as the causative disease mechanism.

Based on this study and evidence from others, including those from other rare disease fields, we suggest that trio-based sequencing should be part of the routine evaluation of patients with a sporadic IEI phenotype (Box 1). An exome-wide analysis should be conducted to identify potentially novel disease genes in cases with a negative diagnostic WES result in whom a strong clinical suspicion for an underlying monogenic cause remains. Thus far, the relative proportion of DNVs among IEI patients with a genetic diagnosis, estimated to be around 6-14%, seems modest compared to other rare disease fields (i.e., >80% in neurodevelopmental disorders (NDDs)) (79). There are several explanations for this difference that suggest that the true contribution of DNVs is higher than currently appreciated. Most importantly, much more experience has been gained with DNV assessment in the field of NDDs. Despite a steep increase in the total diagnostic rate (6, 80, 81) and the identification of 285 developmental disorder (DD)-associated DNVs, modelling suggests that more than 1000 DD-associated genes still remain to be discovered (8). As more trio-based sequencing data will be generated from suspected IEI patients, the field should undertake larger-scale analyses that leverage existing statistical models from the field of NDDs/DDs, including models for gene/exon level enrichment and the identification of gain-of-function nucleotide clusters (8). Moreover, there is still a bias towards autosomal recessive (AR) disease genes in the IEI field, while this imbalance is shifting with the discovery of an increasing number of autosomal dominant (AD) disease genes (74). Trio-based sequencing could accelerate the discovery of mutations in novel AD IEI genes.

### Box 1. Proposed indications for trio-based sequencing in patients with inborn errors of immunity

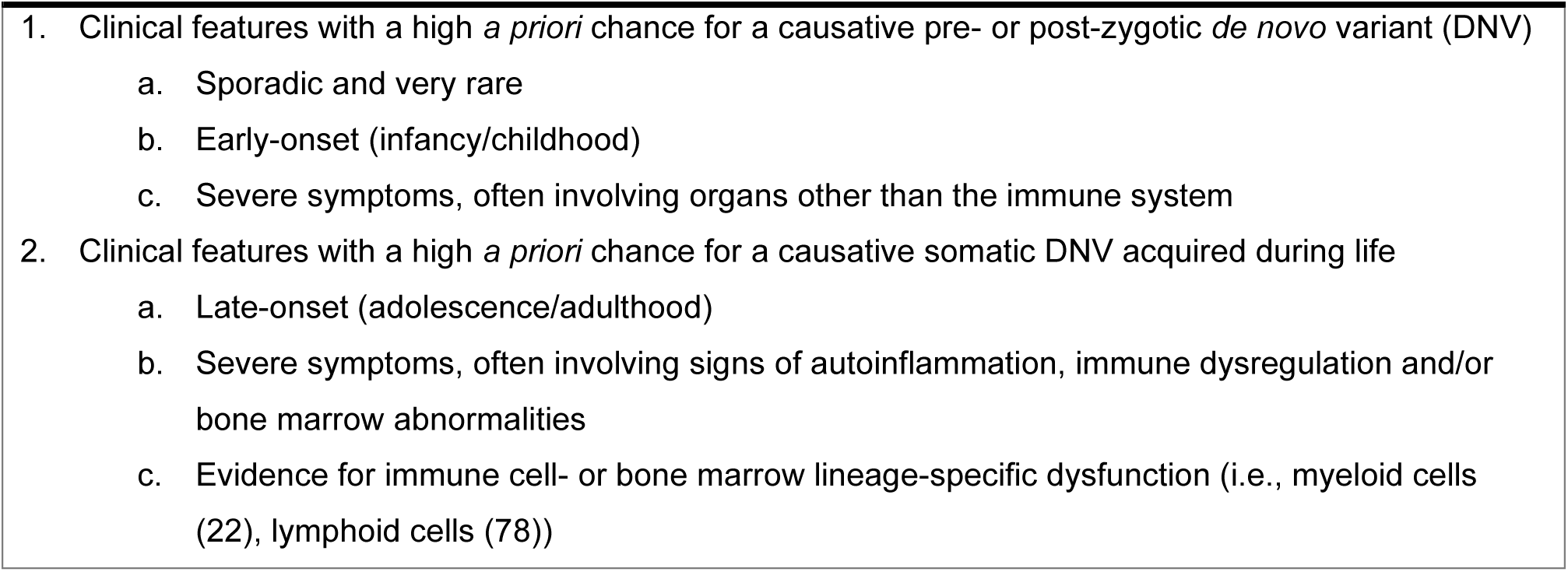

Inborn errors of immunity constitute a large group of heterogeneous disorders with differences in the expected contribution of DNVs. The *a priori* probability for the identification of a DNV will be highest in patients with early-onset, severe phenotypes, such as the combined immunodeficiencies (CID), especially CIDs with syndromic features, and patients with autoinflammatory syndromes and/or immune dysregulation with autoimmunity (Box 1). Although most of the reported genes underlying CIDs follow AR inheritance patterns, many genes following AD and X-linked (dominant) inheritance patterns have been described in recent years (48). The genes affected in these disorders possess high intolerance for loss-of-function mutations and essential biological functions. As expected, the DNVs reported in this category to date act through mechanisms of haploinsufficiency (i.e. *RELA*, pLI 0.999), dominant-negative interference (i.e. *IKZF1*, pLI 0.999 (82); *STAT3*, pLI 1.000 (83)) or complete deficiency in hemizygotic males (i.e. *WAS*, pLI 0.999 (84); *IL2RG*, pLI 0.992 (85)). Some heterozygous DNVs can also cause CID through hypermorphic effects at protein level (*i.e., RAC2*, pLI 0.966 (86)). Trio-based sequencing should also be considered in patients with sporadic autoinflammatory syndromes and/or autoimmunity, even when presenting at an adult age that could suggest somatic *de novo* mutations. In these patients, various pathogenic DNVs in different genes have already been described, originating both from the germline (*PLCG2, STAT1*) and soma (i.e., *NLRP3, UBA1, TLR8*) (13, 18, 21–24). These genes do not necessarily have high constraint for loss-of-function mutations, but they possess nucleotide clusters that are highly conserved and intolerant to variation, encoding protein domains with important regulatory functions.

This explorative study has a number of limitations. First, the sample size precludes a reliable estimation of the prevalence of DNVs among patients with sporadic IEIs. Furthermore, the strict diagnostic rate of both inherited variants and (likely) pathogenic DNVs in our cohort is limited (n=17, 13.8%) compared to other studies. It has been previously reported that the diagnostic yield of WES for IEI patients varies widely from 10-79% (15). The most likely explanation for a relatively low diagnostic yield in our study is the patient selection. We excluded patients with suspected inherited disease but chose not to apply any other selection criteria in order to study a representative cross-section of suspected IEI patients in our centre in whom WES was performed. As a result, patients were included even if the *a priori* chance of an IEI was low but should be ruled out in the differential diagnosis (i.e., new-born screening shows low T cell receptor excision circles (TRECs)). Moreover, compared to other cohorts, the percentage of patients with syndromal CIDs, autoinflammatory syndromes and immune dysregulation was relatively high and could influence the generalisability of our results. Lastly, the functional effect of most candidate DNVs were not evaluated. As DNVs have a high chance of being deleterious, functional experiments should always be attempted to validate the predicted effect. The candidate DNVs in potentially novel IEI genes were shared on GeneMatcher in order to find similar cases that could motivate further investigation into the underlying mechanisms (1, 87).

In conclusion, we applied trio-based whole exome sequencing in a retrospective cohort of 123 patients with sporadic, suspected IEI, leading to the identification of 14 DNVs with a possible or likely chance of pathogenicity. Amongst the candidate DNVs in potentially novel IEI genes, additional functional evidence was provided in support of a pathogenic role for the DNV in *FBXW11* in a patient with an autoinflammatory phenotype. We advocate the structural implementation of trio-based sequencing in the diagnostic evaluation of patients with sporadic IEI. With decreasing costs of exome sequencing, this approach could improve the diagnostic rate of IEI and advance IEI gene discovery.

## Data Availability

DNA sequencing data of patients are not publicly available as it is confidential human subject data. Data supporting the findings of this study are available within the article and its supplementary materials.

## Acknowledgements

We thank the Bioinformatics group of the Genome Diagnostics division of the department of Human Genetics and the Radboud Genomics Technology Center of the Radboud University Medical Center for the sharing, annotation and pseudonymisation of whole exome sequencing datasets of patients and their parents included in this study. Furthermore, we acknowledge all members of the multidisciplinary immunogenetics sign-out meeting of the University Medical Centers from Nijmegen and Maastricht. The authors of this review also acknowledge support from several funding parties. M. G. Netea was supported by an ERC Advanced Grant (No. 833247) and a Spinoza Grant of the Netherlands Organization for Scientific Support. This research was also part of a Radboud Institute for Molecular Life Sciences PhD grant (to M. G. Netea). F. L. van de Veerdonk was supported by a ZonMW Vidi grant and HDM-FUN EU H2020. A. Hoischen was supported by the Solve-RD project of the European Union’s Horizon 2020 research and innovation programme (No. 779257).

## Figure and Table supplements

**Figure 1 – table supplement 1.** List of 123 patient-parent trios with whole exome sequencing performance statistics and associated clinical phenotypes. Patient characteristics, including sex, five-year age range at sampling and phenotypical IUIS classification were provided, and information on the genetic ancestry, median target coverage, % targets covered with at least 5x or 20x coverage was retrieved from each trio-based whole exome sequencing dataset. Patients included in previous publications are noted.

| Patient Nr | Sex | Age range at sampling | Genetic ancestry | Phenotype (IUIS classification) | Median target coverage | % Targets ≥5x | % Targets ≥20x | Previously published |
| --- | --- | --- | --- | --- | --- | --- | --- | --- |
| 1 | M | 11-15 | Europe | SCID | 112 | 98.68 | 93.73 | Yes (28) |
| 2 | M | 16-20 | Europe | Bone marrow failure | 110 | 99.74 | 97.96 | Yes (10) |
| 3 | F | 31-35 | Europe | Immune dysregulation, syndromes with autoimmunity and others | 116 | 99.63 | 98.29 | No |
| 4 | M | 0-5 | Europe | Defect in intrinsic and innate immunity, bacterial and parasitic | 153 | 99.63 | 98.2 | Yes (10) |
| 5 | M | 0-5 | Europe | Predominantly antibody deficiency, hypogammaglobulinemia | 121 | 99.68 | 98.18 | No |
| 6 | M | 21-25 | Europe | Bone marrow failure | 134 | 99.2 | 95.08 | No |
| 7 | M | 46-50 | Europe | Autoinflammatory disorder | 100 | 99.66 | 97.88 | Yes (10) |
| 8 | M | 0-5 | Europe | CID, syndromal | 97 | 99.31 | 96.27 | Yes (10) |
| 9 | M | 6-10 | Europe | Predominantly antibody deficiency, hypogammaglobulinemia | 90 | 99.61 | 97.32 | Yes (10) |
| 10 | F | 0-5 | Europe | Immune dysregulation, HLH/EBV | 112 | 99.63 | 98.3 | Yes (10) |
| 11 | M | 0-5 | Europe | Immune dysregulation, HLH/EBV | 104 | 99.54 | 97.19 | Yes (10) |
| 12 | F | 11-15 | Middle East | CID, syndromal | 106 | 99.61 | 97.8 | Yes (10) |
| 13 | M | 16-20 | Europe | CID, syndromal | 139 | 99.54 | 97.56 | Yes (10) |
| 14 | M | 36-40 | Europe | Unclassified | 240 | 99.37 | 97.66 | Yes (10) |
| 15 | F | 0-5 | Europe | SCID | 86 | 98.8 | 93.7 | Yes (10) |
| 16 | F | 0-5 | Europe | Defect in intrinsic and innate immunity, MSMD and viral infection | 79 | 99.25 | 95.34 | Yes (10) |
| 17 | F | 6-10 | Europe | CID, syndromal | 109 | 99.53 | 97.59 | Yes (10) |
| 18 | F | 26-30 | Europe | CID, syndromal | 117 | 98.61 | 94.32 | Yes (10) |
| 19 | F | 6-10 | Europe | Autoinflammatory disorder | 119 | 99.2 | 96.11 | No |
| 20 | F | 26-30 | No Genetic Ancestry Info | Unclassified | 120 | 99.5 | 97.55 | No |
| 21 | M | 6-10 | Europe | Autoinflammatory disorder | 131 | 99.76 | 98.47 | Yes (10) |
| 22 | F | 31-35 | Europe | Immune dysregulation, autoimmunity and others | 118 | 99.4 | 97.06 | No |
| 23 | M | 11-15 | Middle East | Autoinflammatory disorder | 123 | 99.7 | 98.21 | Yes (10) |
| 24 | M | 0-5 | Europe | CID, syndromal | 119 | 99.4 | 95.92 | Yes (10) |
| 25 | F | 6-10 | Europe | Autoinflammatory disorder | 129 | 99.48 | 97.61 | Yes (10) |
| 26 | F | 0-5 | No Genetic Ancestry Info | Bone marrow failure | 174 | 99.16 | 95.82 | Yes (10) |
| 27 | M | 11-15 | Europe | Autoinflammatory disorder | 144 | 99.35 | 95.85 | No |
| 28 | F | 16-20 | Europe | Predominantly antibody deficiency, hypogammaglobulinemia | 110 | 99.48 | 97.15 | No |
| 29 | F | 26-30 | Europe | Autoinflammatory disorder | 110 | 99.3 | 96.44 | No |
| 30 | M | 6-10 | Europe | Defect in intrinsic and innate immunity, bacterial and parasitic | 102 | 99.27 | 95.47 | No |
| 31 | M | 6-10 | Africa | CID, syndromal | 108 | 99.51 | 96.61 | No |
| 32 | M | 11-15 | Europe | CID, syndromal | 127 | 99.55 | 97.28 | No |
| 33 | F | 36-40 | Europe | Immune dysregulation, autoimmunity and others | 115 | 99.35 | 96.79 | No |
| 34 | M | 0-5 | Europe | Defect in intrinsic and innate immunity, MSMD and viral infection | 119 | 99.15 | 94.49 | No |
| 35 | M | 11-15 | Europe | Defects in intrinsic and innate immunity | 114 | 99.2 | 94.45 | No |
| 36 | M | 0-5 | Europe | Bone marrow failure | 117 | 99.38 | 96.49 | No |
| 37 | M | 6-10 | Europe | Autoinflammatory disorder | 109 | 99.23 | 94.75 | No |
| 38 | F | 16-20 | Europe | Autoinflammatory disorder | 107 | 99.62 | 96.51 | No |
| 39 | F | 31-35 | Europe | Predominantly antibody deficiency, hypogammaglobulinemia | 134 | 99.33 | 96.83 | No |
| 40 | M | 11-15 | Europe | CID, syndromal | 121 | 99.42 | 96.54 | No |
| 41 | F | 51-55 | Europe | Bone marrow failure | 90 | 99.16 | 95.4 | No |
| 42 | M | 0-5 | Europe | Immune dysregulation, HLH/EBV | 106 | 99 | 94.14 | No |
| 43 | F | 0-5 | Europe | CID, syndromal | 106 | 98.5 | 92.92 | No |
| 44 | M | 16-20 | Europe | Predominantly antibody deficiency, hypogammaglobulinemia | 112 | 99.51 | 96.92 | No |
| 45 | M | 0-5 | No Genetic Ancestry Info | CID, syndromal | 225 | 99.04 | 95.09 | No |
| 46 | F | 16-20 | No Genetic Ancestry Info | Predominantly antibody deficiency, hypogammaglobulinemia | 180 | 99.18 | 95.25 | No |
| 47 | M | 16-20 | Europe | Predominantly antibody deficiency, hypogammaglobulinemia | 105 | 99.55 | 96.6 | No |
| 48 | M | 26-30 | No Genetic Ancestry Info | Predominantly antibody deficiency, hypogammaglobulinemia | 230 | 99.32 | 96.19 | No |
| 49 | M | 26-30 | Europe | Predominantly antibody deficiency, hypogammaglobulinemia | 125 | 99.7 | 97.52 | No |
| 50 | M | 6-10 | Europe | CID, syndromal | 102 | 99.54 | 96.62 | No |
| 51 | F | 0-5 | Europe | Autoinflammatory disorder | 125 | 99.58 | 97.78 | No |
| 52 | M | 11-15 | Europe | Predominantly antibody deficiency, hypogammaglobulinemia | 106 | 99.48 | 96.39 | No |
| 53 | F | 11-15 | Europe | Autoinflammatory disorder | 119 | 99.56 | 98.04 | No |
| 54 | F | 11-15 | Middle East | CID, syndromal | 117 | 99.51 | 97.48 | No |
| 55 | M | 6-10 | Europe | Predominantly antibody deficiency, hypogammaglobulinemia | 92 | 99.68 | 97.23 | No |
| 56 | F | 21-25 | Europe | Predominantly antibody deficiency, hypogammaglobulinemia | 109 | 99.56 | 97.52 | No |
| 57 | M | 0-5 | Europe | Autoinflammatory disorder | 103 | 99.67 | 97.25 | No |
| 58 | F | 21-25 | Europe | Unclassified | 113 | 99.39 | 96.88 | Yes (27) |
| 59 | M | 6-10 | Europe | Autoinflammatory disorder | 101 | 98.59 | 93.02 | No |
| 60 | M | 6-10 | Europe | Defects in intrinsic and innate immunity, MSMD and viral infection | 101 | 98.23 | 91.81 | No |
| 61 | M | 0-5 | No Genetic Ancestry Info | CID, syndromal | 21 | 86.15 | 48.53 | No |
| 62 | M | 0-5 | Europe | Immune dysregulation, autoimmunity and others | 125 | 98.93 | 95.02 | No |
| 63 | M | 6-10 | Europe | Immune dysregulation, autoimmunity and others | 105 | 98.63 | 93.33 | No |
| 64 | M | 0-5 | Europe | Immune dysregulation, autoimmunity and others | 120 | 98.9 | 94.92 | No |
| 65 | M | 0-5 | Europe | Defects in intrinsic and innate immunity | 105 | 98.53 | 93.02 | No |
| 66 | F | 0-5 | No Genetic Ancestry Info | CID, syndromal | 240 | 99.24 | 96.69 | No |
| 67 | M | 0-5 | Europe | Autoinflammatory disorder | 104 | 99.07 | 94.62 | No |
| 68 | F | 0-5 | Europe | Defects in intrinsic and innate immunity | 96 | 99.77 | 97.87 | No |
| 69 | M | 0-5 | Europe | CID, non-syndromal | 101 | 99.75 | 98.04 | No |
| 70 | F | 11-15 | Europe | Defects in intrinsic and innate immunity, MSMD and viral infection | 106 | 99.6 | 98.26 | No |
| 71 | F | 6-10 | Europe | Bone marrow failure | 105 | 99.63 | 98.34 | No |
| 72 | M | 0-5 | No Genetic Ancestry Info | Suspected SCID (low TRECs) | 148 | 99.42 | 96.14 | No |
| 73 | M | 0-5 | No Genetic Ancestry Info | Immune dysregulation, autoimmunity and others | 253 | 99.27 | 96.59 | No |
| 74 | M | 31-35 | Europe | Autoinflammatory disorder | 127 | 99.83 | 98.81 | No |
| 75 | F | 6-10 | Europe | Bone marrow failure | 99 | 99.09 | 98.92 | No |
| 76 | F | 0-5 | Europe | Immune dysregulation, autoimmunity and others | 90 | 99.6 | 97.64 | No |
| 77 | F | 0-5 | Europe | CID, syndromal | 116 | 99.66 | 98.32 | No |
| 78 | F | 6-10 | Europe | CID, syndromal | 111 | 99.64 | 98.1 | No |
| 79 | F | 6-10 | Europe | Congenital defect of phagocyte, neutropenia | 92 | 99.56 | 96.55 | No |
| 80 | F | 0-5 | No Genetic Ancestry Info | Suspected SCID (low TRECs) | 206 | 99.26 | 96.32 | No |
| 81 | F | 0-5 | Europe | Congenital defect of phagocyte, neutropenia | 110 | 99.66 | 98.07 | No |
| 82 | F | 26-30 | Europe | Predominantly antibody deficiency, hypogammaglobulinemia | 103 | 99.72 | 97.88 | No |
| 83 | F | 0-5 | No Genetic Ancestry Info | CID, non-syndromal | 181 | 98.91 | 95.1 | No |
| 84 | F | 0-5 | No Genetic Ancestry Info | Autoinflammatory disorder | 169 | 98.76 | 94.24 | No |
| 85 | F | 16-20 | Europe | Predominantly antibody deficiency, hypogammaglobulinemia | 90 | 99.46 | 96.32 | No |
| 86 | M | 6-10 | Middle East | Defect in intrinsic and innate immunity, MSMD and viral infection | 209 | 99.06 | 95.12 | No |
| 87 | M | 11-15 | Europe | Autoinflammatory disorder | 98 | 99.65 | 95.11 | No |
| 88 | F | 26-30 | Europe | Autoinflammatory disorder | 124 | 99.56 | 98.07 | No |
| 89 | F | 16-20 | Europe | CID, syndromal | 107 | 99.48 | 97.46 | No |
| 90 | M | 11-15 | Europe | Unclassified | 103 | 99.71 | 97.5 | No |
| 91 | F | 0-5 | Europe | Suspected SCID (low TRECs) | 116 | 97.67 | 90.26 | No |
| 92 | F | 0-5 | Europe | Suspected SCID (low TRECs) | 194 | 98.79 | 94.47 | No |
| 93 | F | 26-30 | Europe | Immune dysregulation, autoimmunity and others | 115 | 99.64 | 98.14 | No |
| 94 | F | 41-45 | Europe | Unclassified | 119 | 99.61 | 98 | No |
| 95 | M | 11-15 | Europe | Defects in intrinsic and innate immunity | 107 | 99.75 | 97.48 | No |
| 96 | M | 21-25 | Europe | CID, syndromal | 251 | 99.03 | 95.58 | No |
| 97 | F | 16-20 | Europe | Immune dysregulation, autoimmunity and others | 98 | 99.48 | 96.25 | No |
| 98 | M | 0-5 | Europe | SCID | 199 | 99.02 | 95.31 | No |
| 99 | F | 11-15 | Europe | Bone marrow failure | 179 | 98.55 | 93.77 | No |
| 100 | F | 21-25 | Europe | Autoinflammatory disorder | 111 | 99.46 | 97.78 | No |
| 101 | M | 0-5 | Middle East | Immune dysregulation, HLH/EBV | 128 | 98.73 | 92.71 | No |
| 102 | F | 11-15 | Middle East | Immune dysregulation, autoimmunity and others | 87 | 98.34 | 88.22 | No |
| 103 | F | 0-5 | Europe | Immune dysregulation, HLH/EBV | 121 | 99.51 | 97.3 | No |
| 104 | F | 0-5 | Europe | Defects in intrinsic and innate immunity | 247 | 98.9 | 95.47 | No |
| 105 | M | 31-35 | Europe | Defects in intrinsic and innate immunity, MSMD and viral infection | 111 | 99.27 | 98.79 | No |
| 106 | F | 11-15 | Europe | Immune dysregulation, autoimmunity and others | 111 | 99.53 | 97.71 | No |
| 107 | F | 0-5 | Europe | Defects in intrinsic and innate immunity | 98 | 99.4 | 97.09 | No |
| 108 | M | 21-25 | Europe | Bone marrow failure | 159 | 99.35 | 99.21 | No |
| 109 | M | 31-35 | Europe | Congenital defect of phagocyte, neutropenia | 115 | 99.71 | 97.99 | No |
| 110 | F | 0-5 | Europe | Immune dysregulation, autoimmunity and others | 92 | 99.41 | 96.82 | No |
| 111 | F | 0-5 | Europe | Defect in intrinsic and innate immunity, MSMD and viral infection | 101 | 99.52 | 97.5 | No |
| 112 | F | 6-10 | Europe | CID, syndromal | 101 | 99.51 | 97.52 | No |
| 113 | M | 0-5 | Europe | Autoinflammatory disorder | 96 | 99.62 | 96.94 | No |
| 114 | M | 6-10 | Europe | Immune dysregulation, autoimmunity and others | 114 | 99.33 | 99.16 | No |
| 115 | F | 0-5 | Europe | SCID | 151 | 99.1 | 98.96 | No |
| 116 | M | 0-5 | Central South Asia | Bone marrow failure | 125 | 99.34 | 99.17 | No |
| 117 | M | 0-5 | Europe | Congenital defect of phagocyte, neutropenia | 111 | 99.7 | 97.93 | No |
| 118 | F | 0-5 | Middle East | Immune dysregulation, autoimmunity and others | 86 | 99.11 | 98.82 | No |
| 119 | F | 11-15 | Europe | Autoinflammatory disorder | 120 | 99.57 | 97.98 | No |
| 120 | M | 11-15 | Europe | Congenital defect of phagocyte, functional defects | 167 | 99.37 | 99.23 | No |
| 121 | F | 0-5 | Middle East | Defects in intrinsic and innate immunity | 97 | 99.49 | 97.05 | No |
| 122 | M | 0-5 | Europe | Suspected SCID (low TRECs) | 114 | 99.36 | 99.16 | No |
| 123 | M | 0-5 | Europe | Autoinflammatory disorder | 98 | 99.61 | 96.45 | No |
| **Average** |  |  |  |  | **124.11** | **99.23** | **96.18** |  |

Abbreviations: IUIS = International Union of Immunological Societies; (S)CID = (severe) combined immunodeficiency; HLH = haemophagocytic lymphohistiocytosis; EBV = Epstein-Barr virus; MSMD = Mendelian susceptibility to mycobacterial disease; TREC = T cell receptor excision circle.

**Figure 1 – table supplement 2.**
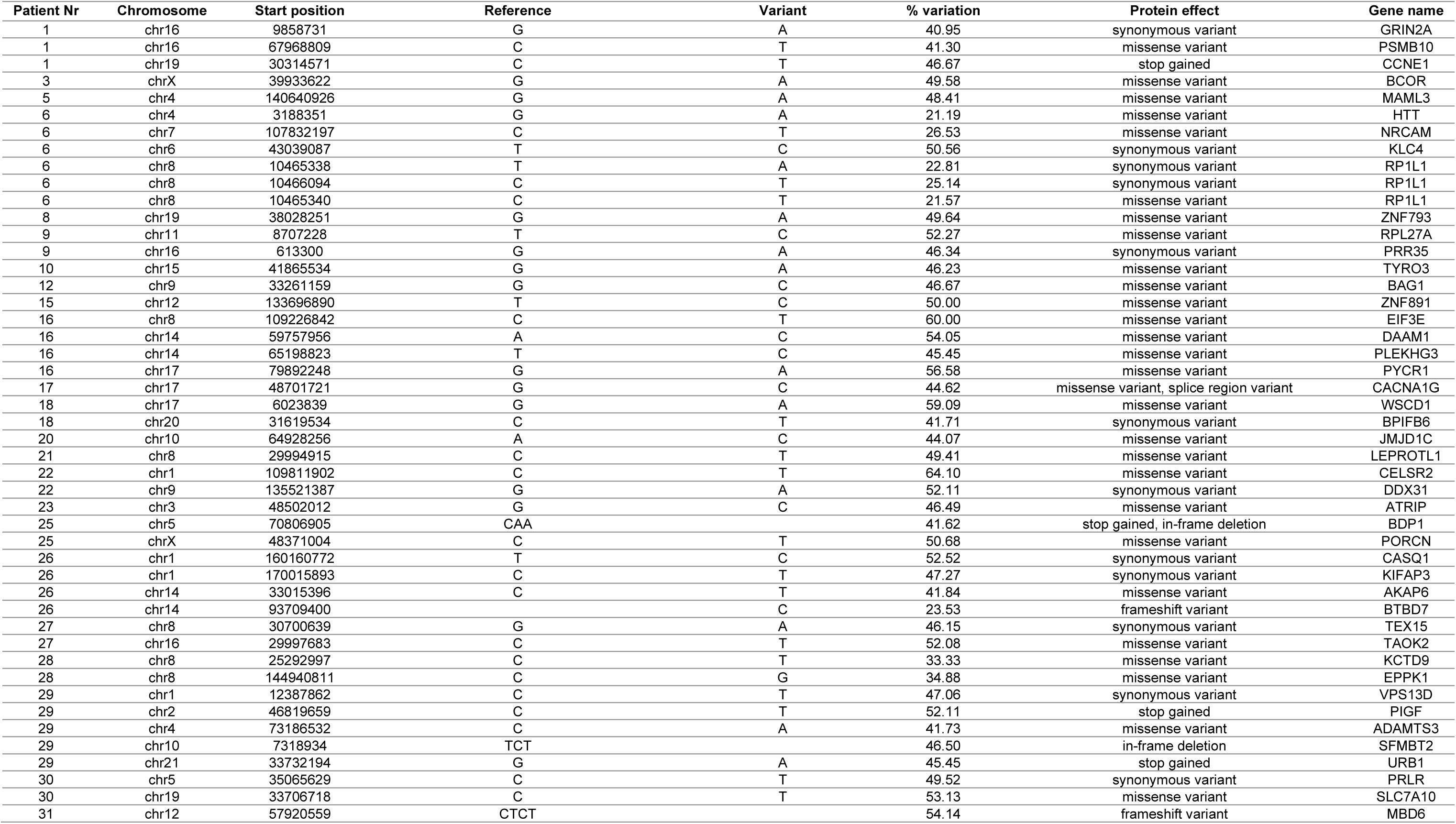
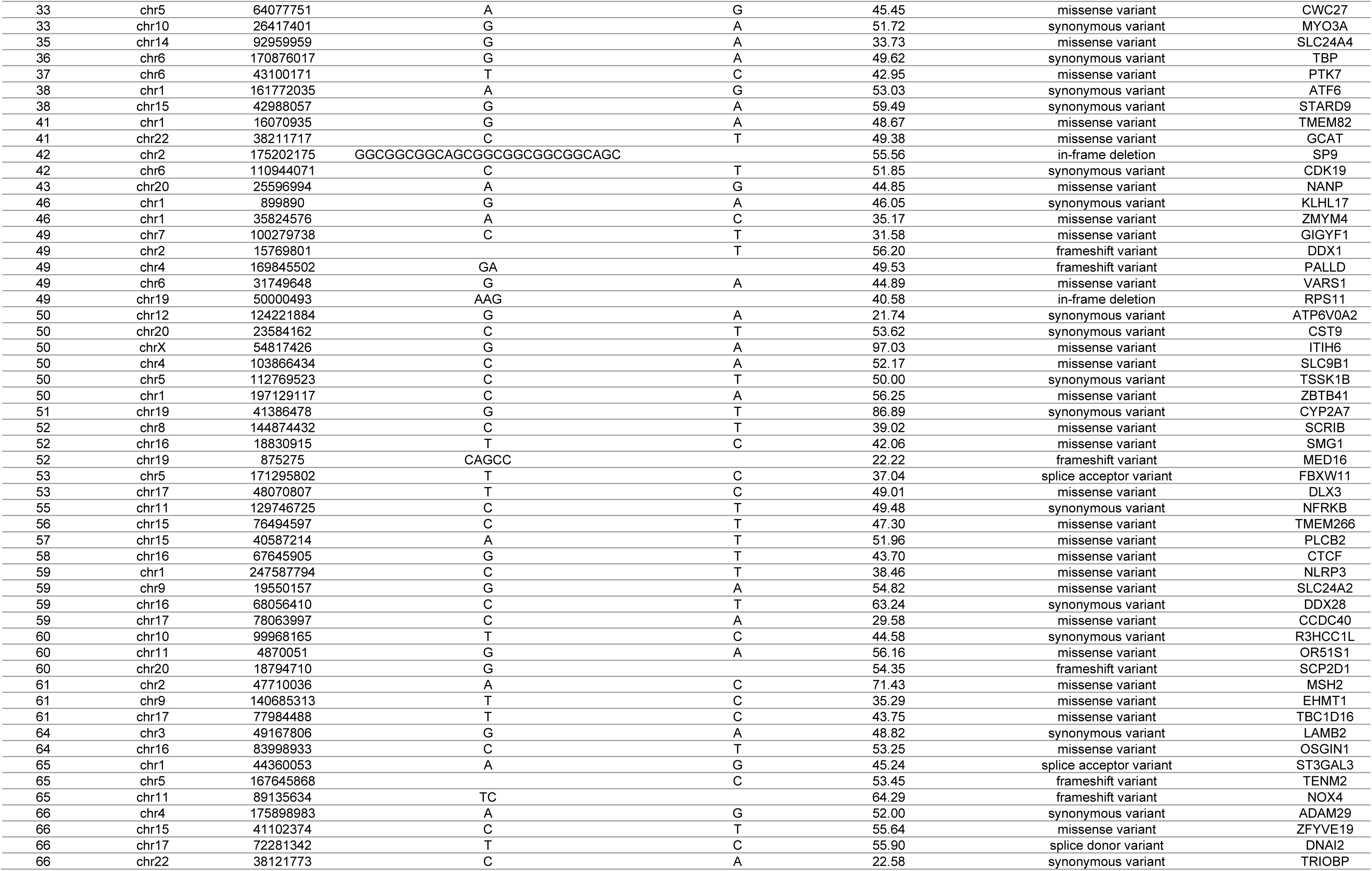
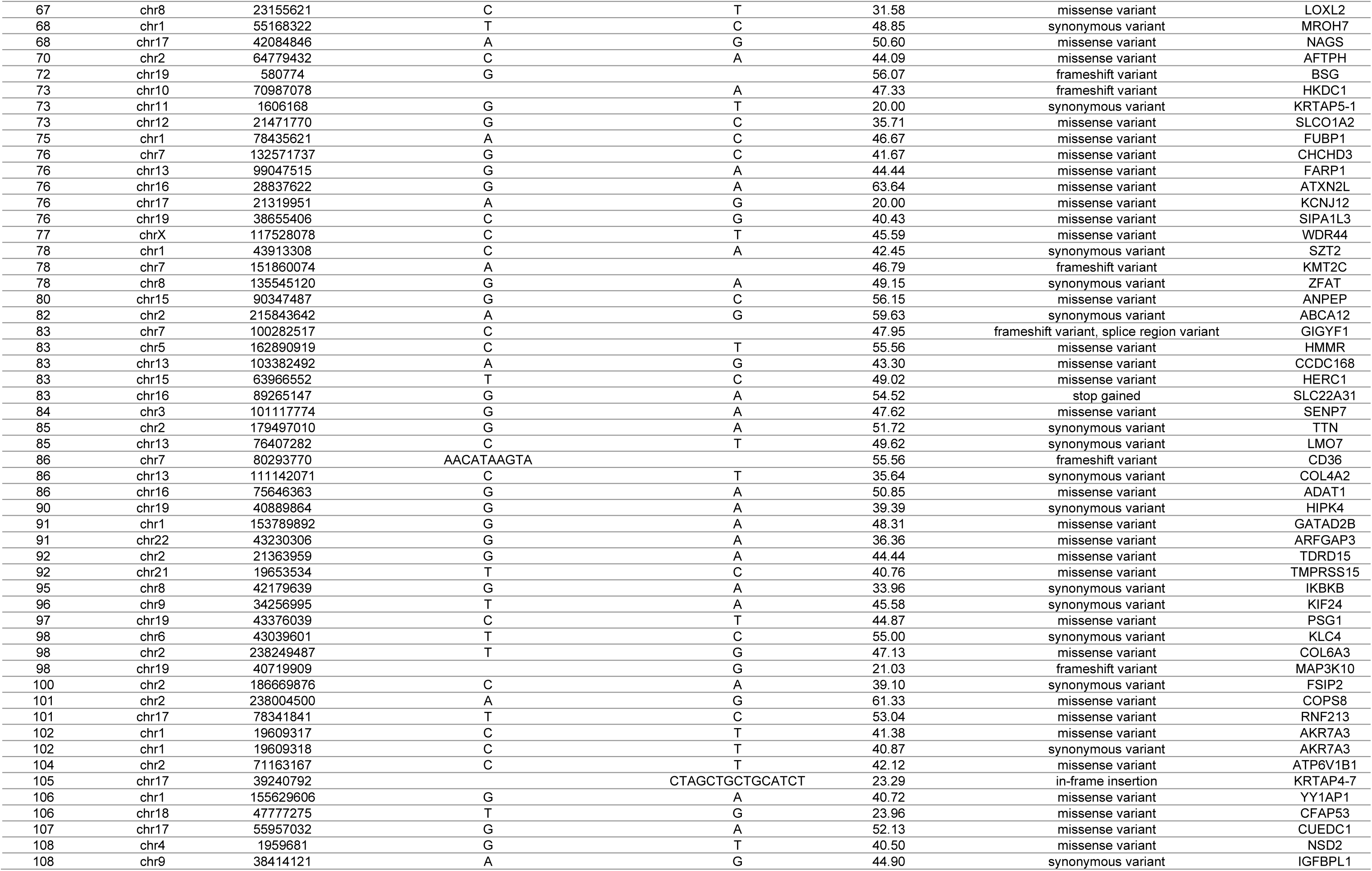
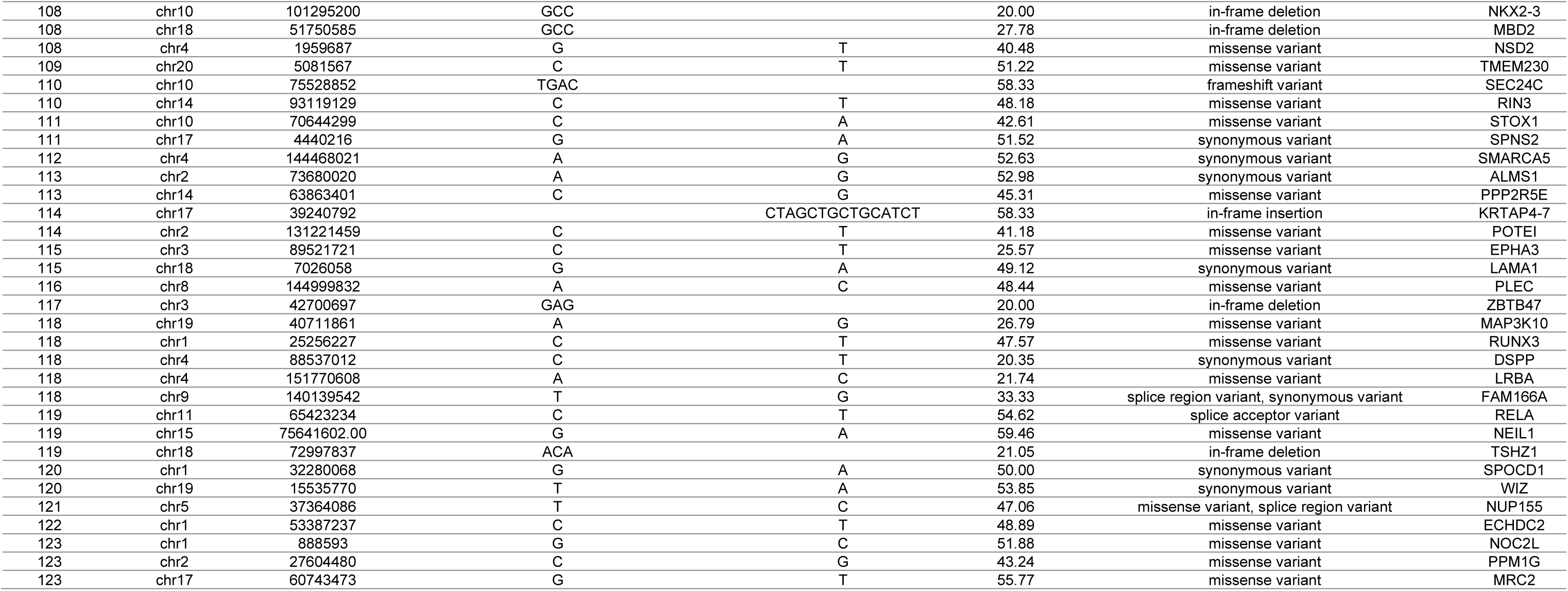
List of all candidate *de novo* variants found in this IEI cohort. Whole exome sequencing datasets of 123 suspected IEI patient-parent trios were consecutively filtered to retain candidate (rare and exonic or splice site region) *de novo* variants of each patient.

**Figure 1 – table supplement 3.**
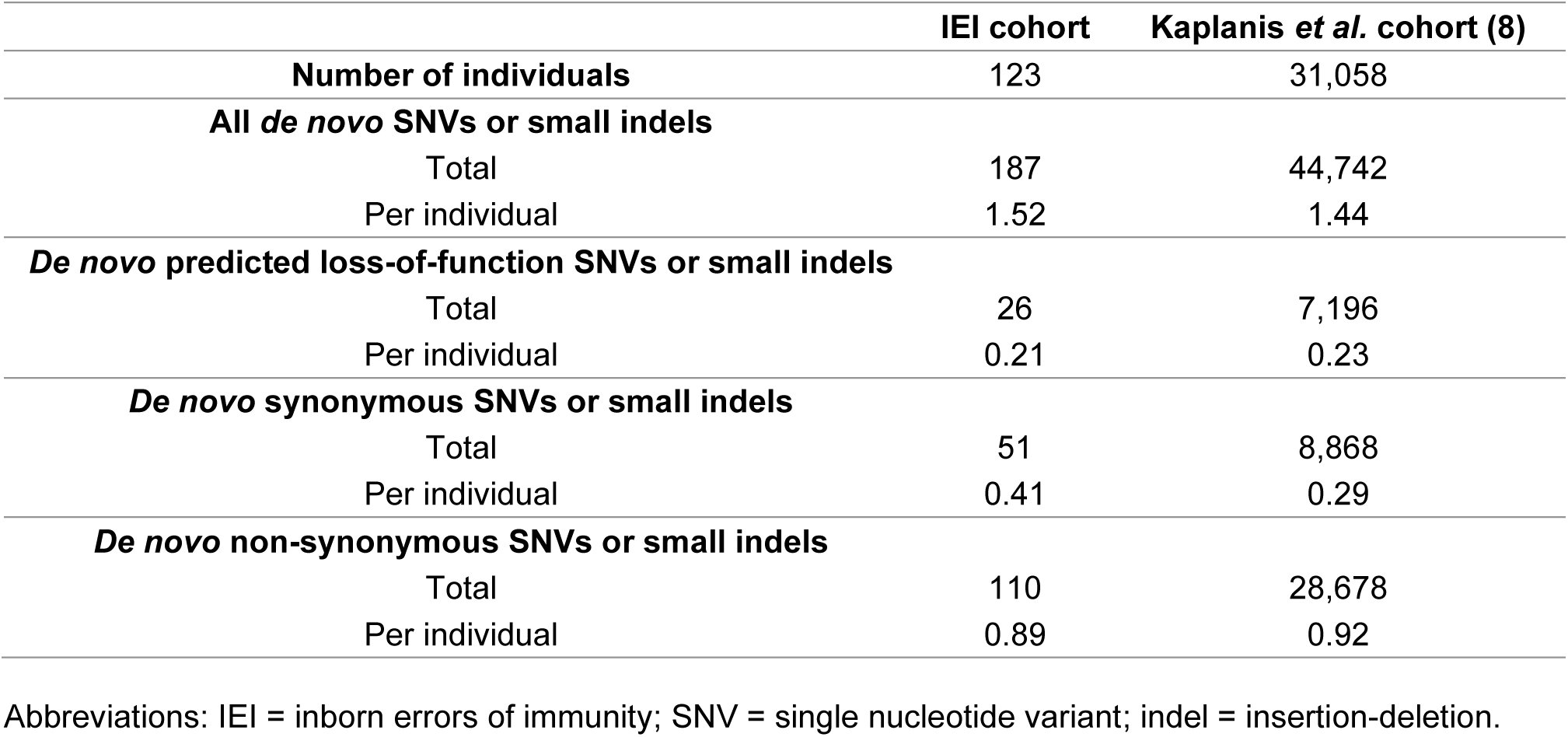
*De novo* variant rate and distribution of *de novo* variant types across our IEI cohort in comparison to a reference cohort from Kaplanis *et al.* (8). The total amount of candidate *de novo* variants was retrieved from our cohort, consisting of 123 individuals. All identified *de novo* variants with minimally 20% variation reads of the comparably processed Kaplanis *et al.* cohort were obtained (8). The amount of predicted loss-of-function, synonymous and non-synonymous *de novo* single nucleotide variants or small insertion-deletions were extracted from both cohorts. The total amount of all, predicted loss-of-function, synonymous and non-synonymous *de novo* variants were divided by the respective cohort size to obtain the average number of respective variants per individual for each cohort.

**Figure 1 – figure supplement 1.**
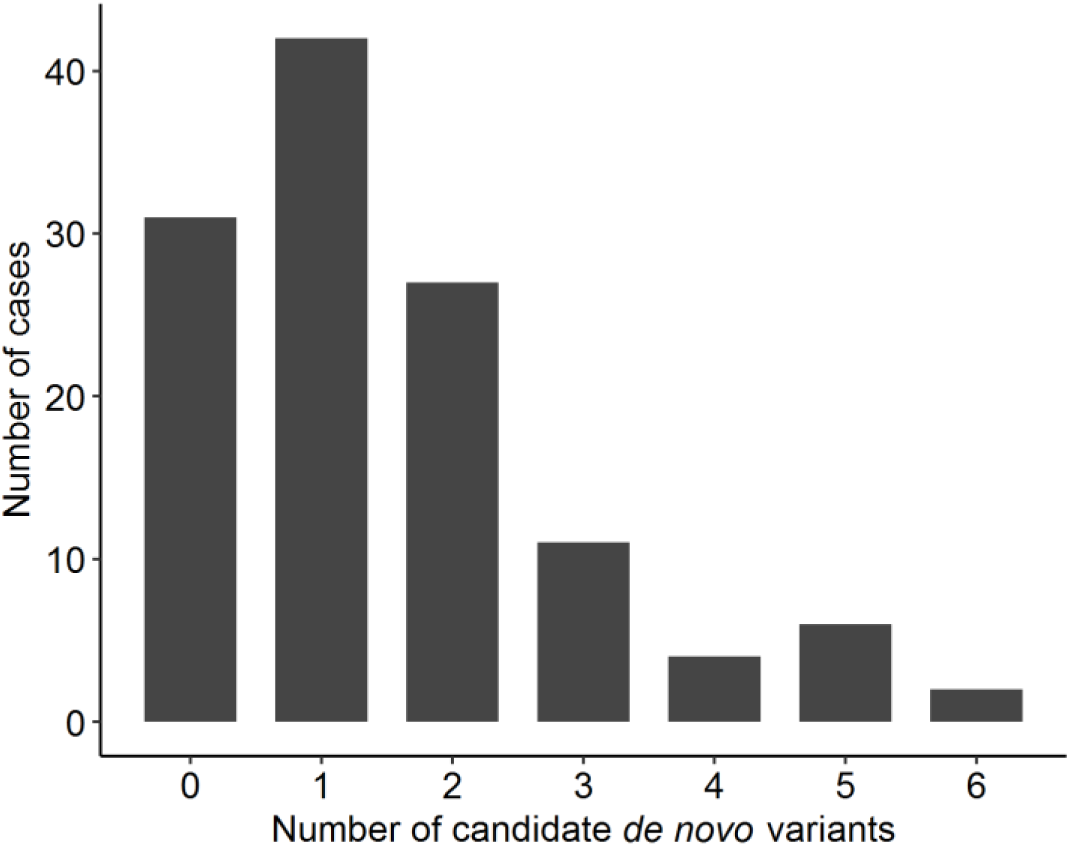
Distribution of *de novo* variants per case. Using the R software, a code was developed that filtered each trio-based whole exome sequencing dataset consecutively on the inheritance, genomic location and population allele frequency of the called single nucleotide variants and small insertion-deletions in order to obtain candidate *de novo* variants. These variants are located in exonic or splice site regions and rarely occur in the population. The number of candidate *de novo* variants were counted per patient.

**Table 1B – table supplement 1.**
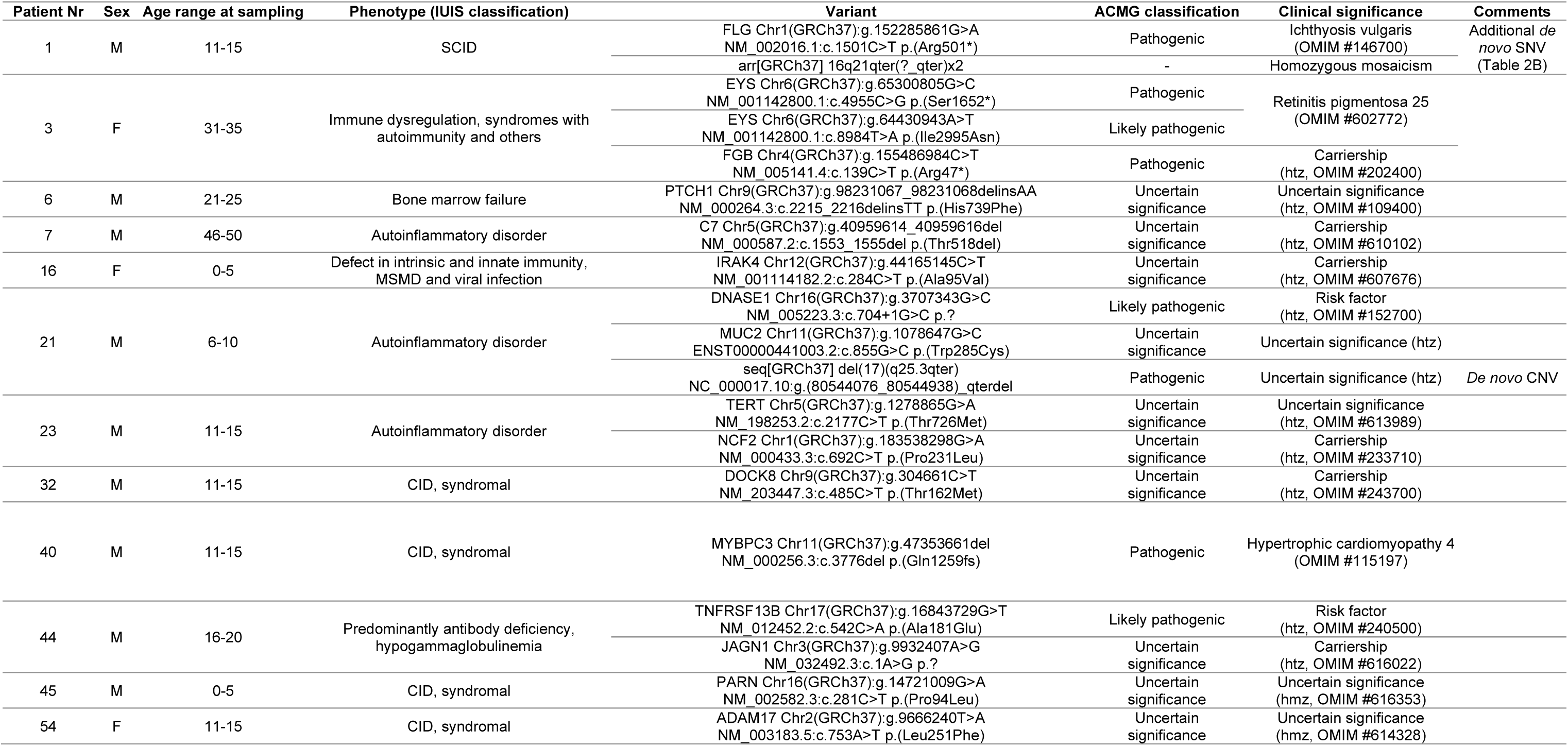
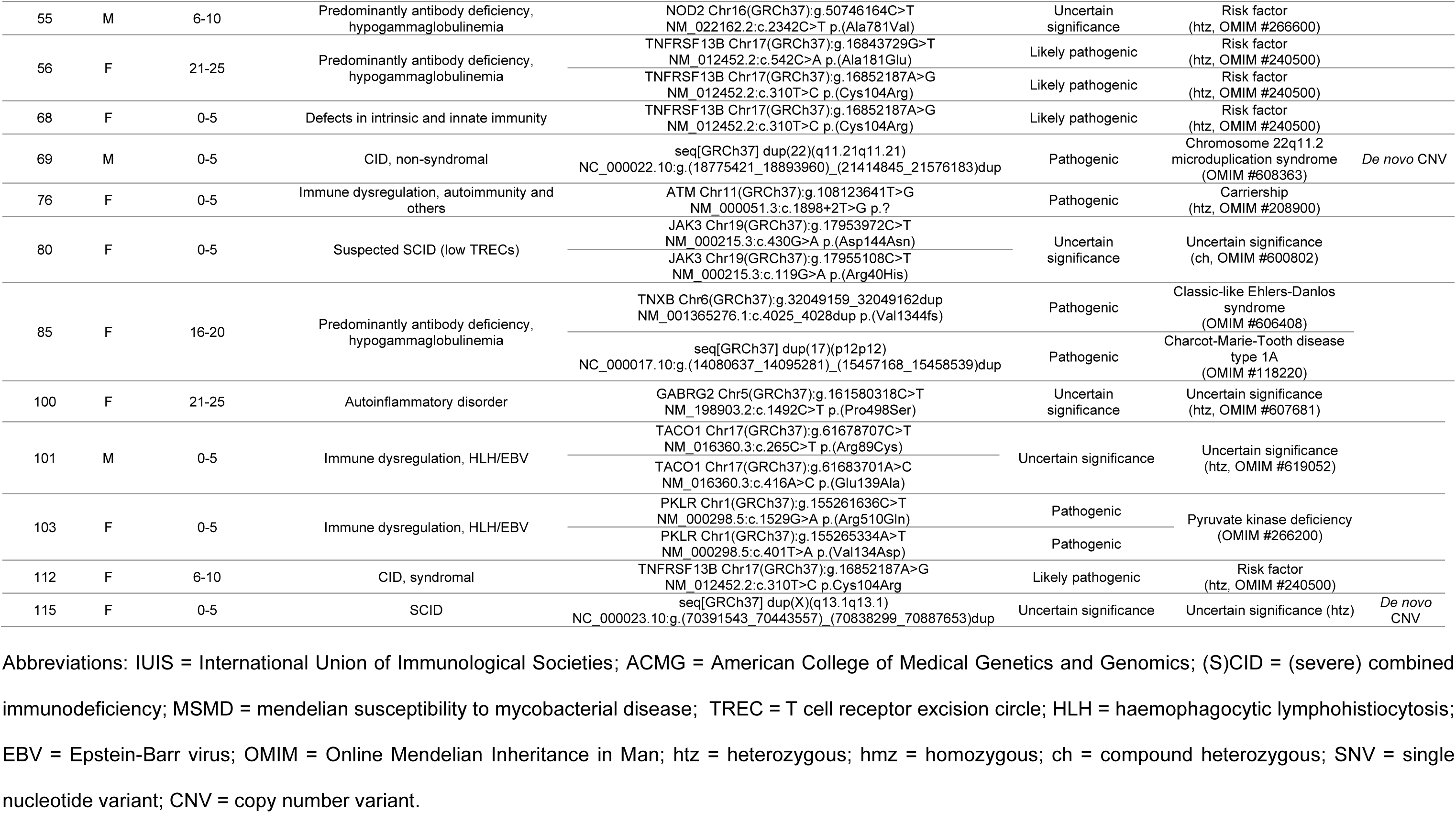
List of patient trios with variants identified in genes outside the diagnostic IEI gene panel, or classified as risk factors, carriership or variants of uncertain significance. Information on copy number variants or inherited single nucleotide variants and small insertion-deletions that were identified prior to this cohort study in the scope of *in silico* gene panel analysis and, if appropriate informed consent was provided, in-depth variant analysis of each patient in this inborn errors of immunity cohort. Variants and their associated American College of Medical Genetics and Genomics classification and clinical significance, including possibly associated clinical phenotypes, are listed.

**Figure 2 – figure supplement 1.**
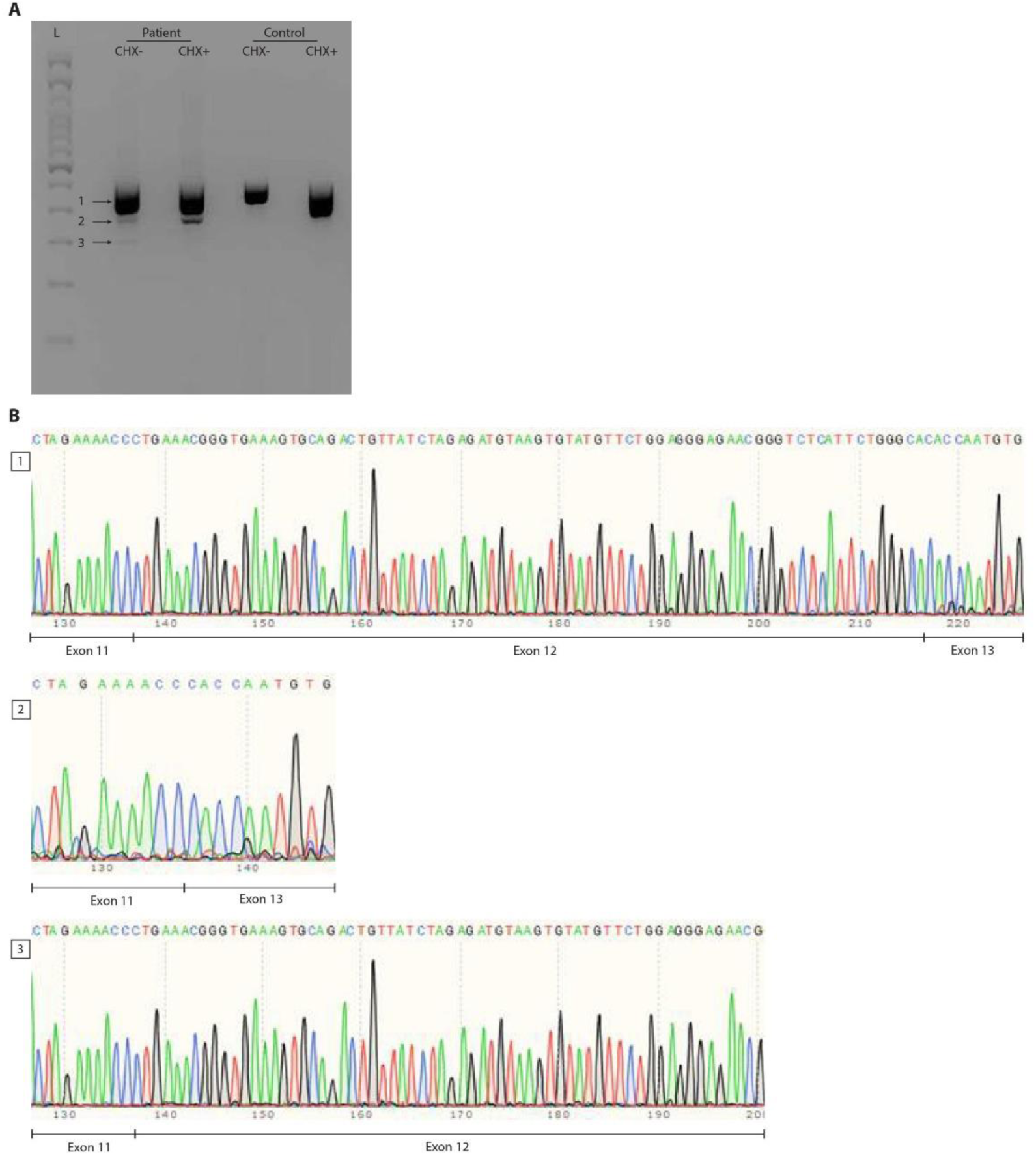
RNA splicing effect of the *FBXW11 de novo* splice site variant (c.1468-2A>G). Panel A shows the agarose gel on the cDNA PCR products of patient and control Epstein–Barr virus (EBV)-transformed lymphoblastoid cell lines (EBV-LCLs) treated with or without cycloheximide (CHX). Three distinct bands were identified and are indicated by arrows next to a 100bp ladder (L). Both the wildtype allele of the patient and the control show a smear, possibly indicating the presence of multiple *FBXW11* isoforms. Panel B shows traces of the three bands from the agarose gel that were cut out and sent for Sanger sequencing. As the splice site variant in the patient was expected to lead to skipping of exon 12, the boundaries between exons 11, 12 and 13 were shown. The second band confirms skipping of exon 12 that results in a shorter transcript of the mutated *FBXW11* allele. After CHX treatment, this band shows an increased quantity, indicating that the mutated allele undergoes nonsense mediated decay, but incomplete. Furthermore, a smaller transcript is formed in the patient, which is shown to contain part of exon 12, but not exon 13. ***Figure 2 – figure supplement 1A – source data 1*.**

